# Repeat polymorphisms in non-coding DNA underlie top genetic risk loci for glaucoma and colorectal cancer

**DOI:** 10.1101/2022.10.11.22280955

**Authors:** Ronen E. Mukamel, Robert E. Handsaker, Maxwell A. Sherman, Alison R. Barton, Margaux L. A. Hujoel, Steven A. McCarroll, Po-Ru Loh

**Affiliations:** Division of Genetics, Department of Medicine, Brigham and Women’s Hospital and Harvard Medical School, Boston, Massachusetts, USA; Center for Data Sciences, Brigham and Women’s Hospital, Boston, Massachusetts, USA; Program in Medical and Population Genetics, Broad Institute of MIT and Harvard, Cambridge, Massachusetts, USA; Stanley Center for Psychiatric Research, Broad Institute of MIT and Harvard, Cambridge, Massachusetts, USA; Department of Genetics, Harvard Medical School, Boston, Massachusetts, USA; Computer Science and Artificial Intelligence Laboratory, Massachusetts Institute of Technology, Cambridge, Massachusetts, USA; Bioinformatics and Integrative Genomics Program, Department of Biomedical Informatics, Harvard Medical School, Boston, Massachusetts, USA

## Abstract

Many regions in the human genome vary in length among individuals due to variable numbers of tandem repeats (VNTRs). We recently showed that protein-coding VNTRs underlie some of the strongest known genetic associations with diverse phenotypes. Here, we assessed the phenotypic impact of VNTRs genome-wide, 99% of which lie in non-coding regions. We applied a statistical imputation approach to estimate the lengths of 9,561 autosomal VNTR loci in 418,136 unrelated UK Biobank participants. Association and statistical fine-mapping analyses identified 107 VNTR-phenotype associations (involving 58 VNTRs) that were assigned a high probability of VNTR causality (PIP≥0.5). Non-coding VNTRs at *TMCO1* and *EIF3H* appeared to generate the largest known contributions of common human genetic variation to risk of glaucoma and colorectal cancer, respectively. Each of these two VNTRs associated with a >2- fold risk range across individuals. These results reveal a substantial and previously unappreciated role of non-coding VNTRs in human health.

## Introduction

Thousands of human genome segments are present in variable numbers of tandem repeats (VNTRs) in different individuals’ genomes, but the effects of VNTRs on human phenotypes have been difficult to measure. At each VNTR locus, a sequence of nucleotides, from seven to thousands of base pairs long, is repeated several to hundreds of times per allele, with the number of repeats varying among individuals. Extreme VNTR alleles have been implicated in human diseases including progressive myoclonus epilepsy (Lalioti et al. 1997) and facioscapulohumeral muscular dystrophy (Wijmenga et al. 1992). However, VNTRs have not been measured in most genome-wide association studies because such polymorphisms are not measured directly by SNP arrays and are challenging to characterize from short sequence reads.

Recent computational advances have enabled VNTR lengths to be measured or estimated from sequencing data and evaluated for association with phenotypes. Most studies to date have analyzed cohorts in which participants are both phenotyped and sequenced, measuring VNTR allele lengths either directly from spanning reads or indirectly from sequencing depth-of-coverage (Course et al. 2020, Bakhtiari et al. 2021, Eslami Rasekh et al. 2021, Garg et al. 2021, Lu et al. 2021, Garg et al. 2022). This approach has succeeded in identifying associations between VNTRs and the expression of nearby genes (Bakhtiari et al. 2021, Eslami Rasekh et al. 2021, Garg et al. 2021, Lu et al. 2021), but discovering associations with health and disease phenotypes (Garg et al. 2022) has proven more difficult due to the challenge of amassing phenotype and VNTR-allele information in the large number of individuals typically needed for genetic studies to discover genotype-phenotype associations, and the still-larger sample sizes required to distinguish among the effects of genomically nearby variants (such as VNTRs and nearby SNPs). An approach that has driven discovery of many SNP-phenotype associations is to impute untyped alleles based on the SNP haplotypes on which they segregate (Marchini et al. 2007); this approach has been extended to complex and multi-allelic copy number variations (Handsaker et al. 2015; Sekar et al. 2016; Boettger et al. 2016). We and others recently observed that this approach can be extended to tandem repeats (Saini et al. 2018, Mukamel et al. 2021, Beyter et al. 2021). We further demonstrated that analysis of shared haplotypes can, at many loci, substantially improve the accuracy of VNTR length estimates from short-read sequencing depth by effectively combining measurements across individuals who inherited identical VNTR alleles from a recent common ancestor (Mukamel et al. 2021).

Our recent work applied this statistical imputation framework to analyze exome-sequencing data in UK Biobank (UKB), showing that protein-coding VNTRs underlie some of the strongest known genetic associations with diverse phenotypes including height, serum urea, and hair curl (Mukamel et al. 2021). Here, we applied this approach to whole-genome sequence data to estimate VNTR lengths genome-wide in UKB participants and assess the role of non-coding as well as coding VNTRs in shaping human phenotypes.

## Results

### Ascertainment and genotyping of 15,653 VNTR polymorphisms genome-wide

We identified VNTR loci across the human genome by analyzing the GRCh38 reference genome in conjunction with 64 haploid genome assemblies generated from long-read sequencing by the Human Genome Structural Variant Consortium (HGSVC2; Ebert et al. 2021). At each of 100,844 autosomal repeats with repeat unit ≥7bp (identified in GRCh38 using Tandem Repeats Finder (TRF); Benson et al. 1999), we determined the lengths of the corresponding repeat alleles in HGSVC2 assemblies by aligning flanking sequences from the human reference (Li 2018), excluding a small fraction of repeats (5.2%) for which either of the two flanking sequences failed to map uniquely in >50% of assemblies (Supplementary Note). Most repeats identified by TRF were either monomorphic (51%) or biallelic (20%) in the HGSVC2 assemblies. Restricting to multiallelic repeats (≥3 distinct alleles) and removing overlapping repeats left 15,653 multiallelic VNTR loci for downstream analysis in whole-genome sequencing (WGS) data (Supplementary Table 1). These VNTRs had a median repeat unit length of 34bp and a median of 6 distinct alleles represented among HGSVC2 assemblies. VNTRs with more repeats generally exhibited greater allelic diversity (Fig. 1a). VNTR allele length distributions in HGSVC2 assemblies had a median range of 199bp and median standard deviation of 46.8bp (Fig. 1b).

**Figure 1.**
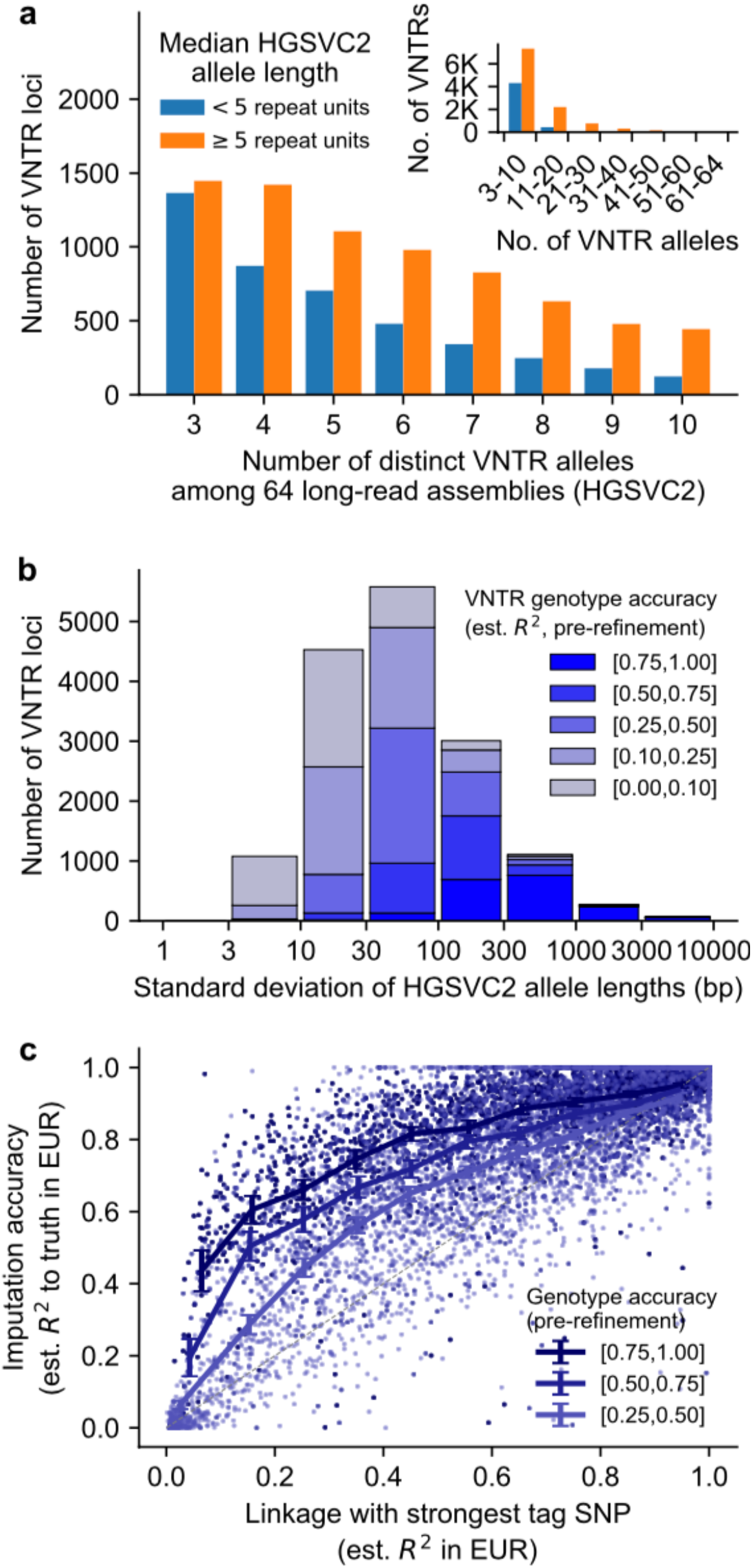
Ascertainment, genotyping and imputation of 15,653 multiallelic VNTR loci. **a)** Counts of VNTR loci stratified by number of distinct alleles observed among *N=*64 long-read haploid genome assemblies from HGSVC2 (x-axis) and the median number of repeats per allele (blue/orange bars). Inset, same counts binned at coarser scale. **b)** Counts of VNTR loci stratified by HGSVC2 allele length distribution width (standard deviation) and estimated accuracy of VNTR genotypes pre-refinement (i.e., measured from WGS depth-of-coverage in individual genomes; Supplementary Note). **c)** Scatter of imputation accuracy vs. level of linkage disequilibrium with the best tag SNP for each VNTR. Color indicates pre-refinement genotype accuracy as in b); VNTRs with noisy estimates of imputation accuracy due to low pre-refinement genotype accuracy (*R*^2^<0.25) were omitted, leaving *N*=7,145 VNTRs for plotting. Lines represent mean imputation accuracy at loci binned by level of linkage with SNPs. Error bars, 95% CIs; EUR, European-ancestry; est., estimated.

To estimate haplotype-resolved VNTR allele lengths in UK Biobank participants, we applied a two-stage approach in which we first generated a reference panel of 9,376 VNTR+SNP haplotypes by analyzing short-read whole-genome sequencing (WGS) data from the Simons Simplex Collection (SSC; Fischbach and Lord 2010, An et al. 2018) and subsequently imputed VNTR alleles from SSC into UKB. To generate the reference panel, we first estimated individual-level VNTR lengths (summed across the two parental alleles) from sequencing depth-of-coverage in 8,936 SSC participants (including 4,688 unrelated individuals whose haplotypes formed the reference panel). Such read-depth-based analysis is capable of distinguishing allele length variation at the scale of hundreds of base pairs (Handsaker et al. 2015); accordingly, these initial VNTR length genotypes captured allelic variation accurately (based on sibling concordance) for highly length-polymorphic VNTRs but less accurately for VNTRs with less-variable lengths (Fig. 1b). To enable analysis of less-length-variable VNTRs and imputation into SNP haplotypes, we next analyzed the SNPs surrounding each VNTR to identify individuals who were likely to have inherited identical VNTR alleles from a recent common ancestor, allowing us to simultaneously reduce noise in VNTR length measurements and estimate haplotype-resolved lengths of individual VNTR alleles. We additionally determined locus-specific parameters for imputing VNTR allele lengths into SNP haplotypes, using cross-validation to assess imputation accuracy and optimize parameters (Mukamel et al. 2021) (Supplementary Note).

For most multiallelic VNTRs, this combination of sequencing read-depth analysis with haplotype-sharing analysis enabled robust statistical imputation that correlated with actual allele-length variation more strongly than any nearby biallelic SNP did (Fig. 1c, Supplementary Table 1, and Supplementary Note) – offering the potential for downstream discovery of genotype-phenotype associations previously invisible to or only weakly discernible from SNP-association analyses. Among 15,653 autosomal, multiallelic repeat loci, this analysis strategy typically captured a substantial proportion of allelic variation (median imputation *R*^2^=0.48), with the most variable VNTRs (allele length s.d. >100bp; 4,462 loci) particularly well-analyzed (median imputation *R*^2^=0.79). Excluding poorly-imputed (*R*^2^<0.1) VNTRs and VNTR regions at which sequencing depth measurements failed quality control filters (Supplementary Note) left 9,561 VNTRs for imputation into UKB.

### Exploring the phenotypic effects of non-coding VNTRs

We applied our statistical imputation framework to estimate allele lengths of 9,561 VNTRs in 418,136 unrelated UKB participants of European ancestry and test these VNTRs for association with 786 phenotypes (including quantitative traits and binary disease phenotypes; Supplementary Table 2), adjusting for age, age^2^, sex, UKB assessment center, genotyping array, and 20 genetic principal components (Methods). These analyses identified 4,968 significant VNTR-phenotype associations (*P* < 5 × 10^−9^), of which 107 associations (involving 58 distinct VNTRs) were assigned a high probability of causality by statistical fine-mapping (FINEMAP posterior inclusion probability (PIP)>0.5 (Benner et al. 2016); Fig. 2, Supplementary Table 3, and Methods). These included five associations between non-coding VNTR polymorphisms and human diseases, including a previously reported association of a VNTR upstream of the insulin gene *INS* with type 1 diabetes, as well as associations of VNTR length polymorphisms with risk of glaucoma, colon polyps, and hypertension that, to our knowledge, have not been previously reported. Two non-coding VNTRs (within *TMCO1* and near *EIF3H*) appeared to generate the largest known contributions of common human genetic variation to risk of glaucoma and colorectal cancer, respectively. The remaining 102 associations involved quantitative traits. Several non-coding VNTRs including a large intronic repeat in *CUL4A* associated strongly with blood cell traits (*P* < 10^−50^), with association strengths similar to those we recently observed for coding VNTRs (Fig. 2). We also tested each VNTR for association with autism in SSC, but did not observe any associations that reached our significance threshold (*P*<5 × 10^−9^).

**Figure 2.**
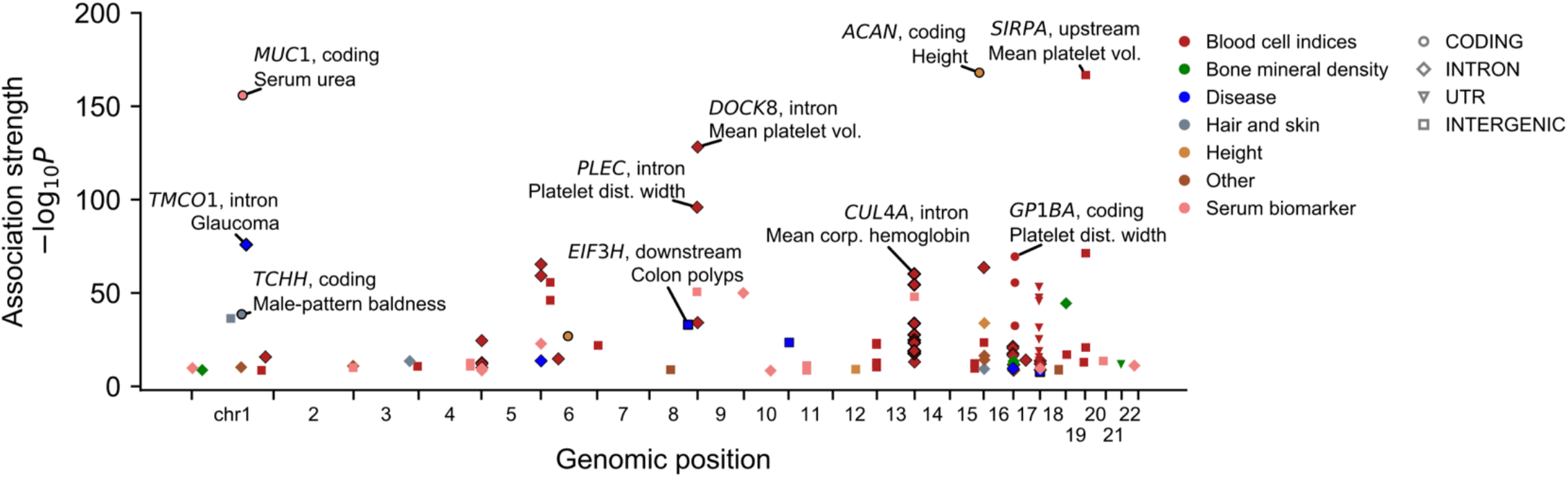
Phenome-wide association and statistical fine-mapping analyses identify 58 VNTRs linked to complex traits. Manhattan plot displaying 107 VNTR-phenotype associations (involving 58 distinct VNTRs) that reached Bonferroni significance (*P*<5 × 10^−9^) and for which the VNTR was assigned a high posterior probability of causality by FINEMAP (PIP>0.5). Marker color indicates phenotype category, and marker shape indicates genic context. Outlined markers indicate associations for which we improved VNTR genotyping or refined the associated phenotype (Supplementary Table 4). For context, the plot also includes two associations to protein-coding VNTRs (at *MUC1* and *TENT5A*, Mukamel et al. 2021) that we previously identified in analysis of whole-exome sequencing data.

For three VNTRs with particularly strong and interesting phenotype associations—at *TMCO1, EIF3H*, and *CUL4A—*we performed a rigorous suite of follow-up analyses that confirmed the robustness and further elucidated the nature of their phenotype associations. First, we improved accuracy with which VNTR repeat numbers could be inferred from WGS (and directly validated this approach using HGSVC2 long-read assemblies; Supplementary Fig. 1) by leveraging subsequent whole-genome sequencing of 200,018 UKB participants and developing statistical models tailored to the allele distribution at each locus (Supplementary Note). In each case, the absolute strength of the VNTR’s association with phenotype, as well as its strength relative to nearby SNP associations, increased with this improved analysis (Supplementary Table 4). We also refined definitions of disease phenotypes in UKB (Supplementary Note) and analyzed data from independent cohorts to replicate associations and search for insights into potential molecular mechanisms as detailed below.

### Repeat expansion at *TMCO1* associates with glaucoma risk more strongly than any SNP or indel in the genome

The strongest disease association we observed involved expansion into many repeats of an intronic 28bp sequence in *TMCO1*; this VNTR associated with glaucoma risk more strongly than any SNP or indel in the entire genome (*P=*1.3 × 10^−76^ vs. 2.8 × 10^−68^ for the strongest SNP association genome-wide; Fig. 3a,b). All the expanded VNTR alleles (containing 5-11 repeat units vs. the one-repeat major allele; Fig. 3a) segregated on a common ∼70kb SNP haplotype at *TMCO1* (AF=12% in UKB) that was among the first-identified and strongest known influences of common genetic variation on glaucoma (Burdon et al. 2011) (Fig. 3b); in our analysis, excess cases among carriers of expanded alleles accounted for ∼10% of primary open-angle glaucoma cases in UKB. Glaucoma is the leading cause of irreversible blindness worldwide (Steinmetz et al. 2021), characterized by optic nerve damage caused in most cases by elevated intraocular pressure (IOP). Even after excluding glaucoma cases, *TMCO1* VNTR length associated with IOP more strongly than any SNP in the genome did, providing independent statistical evidence that the VNTR rather than nearby SNPs underlies the GWAS signal at *TMCO1* (*P*=6.5 × 10^−60^vs. *P*>2.9 × 10^−51^ for SNPs in analyses of *N*=94,877 UKB participants with IOP phenotypes and no reported glaucoma; Fig. 3c), and that the VNTR affects glaucoma risk through its effect on IOP.

**Figure 3.**
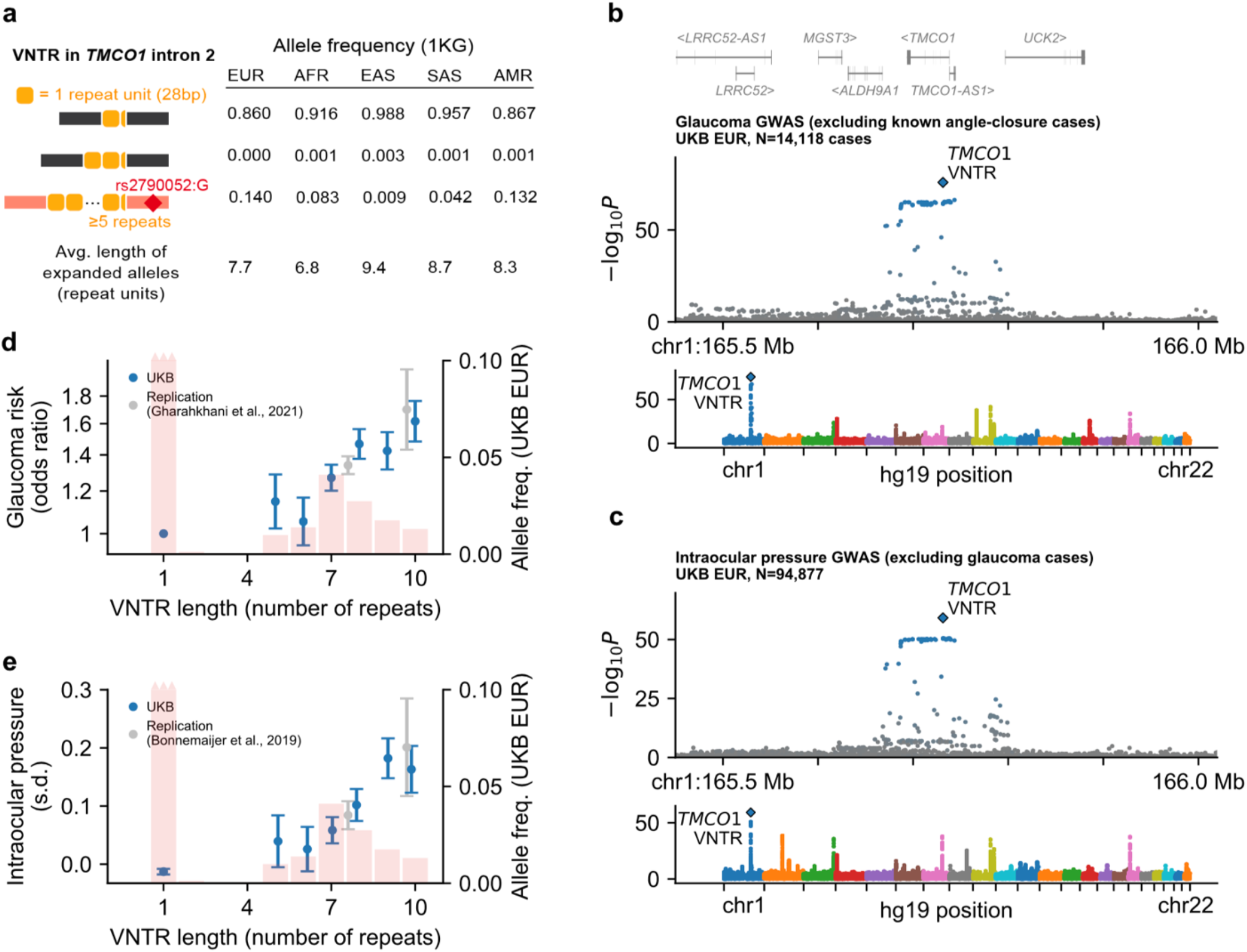
An intronic repeat expansion within *TMCO1* associates with glaucoma risk and intraocular pressure. **a)** Frequencies of the 1-, 2-, and ≥5 repeat unit alleles in each of the continental populations represented in the 1000 Genomes Project. Expanded alleles (5-11 repeat units) segregated with a ∼70kb SNP haplotype (red) represented by rs2790052:G. Each allele in HGSVC2 also contains a partial repeat (7bp of the 28bp unit) depicted in the haplotype diagrams. **b,c)** Associations of SNPs and VNTR with glaucoma (b) and intraocular pressure (c). SNP and VNTR associations are shown at the *TMCO1* locus (top) and genome-wide (bottom). Colored markers in locus plots, variants in partial LD with the VNTR (*R*^2^>0.01). **d,e)** Effect sizes of VNTR alleles for glaucoma risk (d, left axis) and mean intraocular pressure in carriers of each allele (e, left axis). Values in UK Biobank are shown in blue; values inferred based on SNP associations in independent replication cohorts are shown in gray (Supplementary Note). Histograms (right axis), frequencies of VNTR allele lengths estimated in European-ancestry UKB participants. Error bars, 95% CIs.

Repeat alleles of the *TMCO1* VNTR formed an allelic series with increasing effects on IOP and glaucoma risk at longer repeat lengths (Fig. 3d,e). The longest VNTR alleles (top 3%) associated with larger effects on IOP and glaucoma risk than any common SNP elsewhere in the genome (glaucoma OR=1.51 [95% CI, 1.42–1.60] vs. OR≤1.34 for unlinked SNPs with MAF>0.01; IOP β=0.185 s.d. (SE, 0.013 s.d.) vs. β≤0.155 s.d. for SNPs). Individuals homozygous for long alleles (top 0.3% of summed allele length) exhibited >2-fold increased glaucoma risk relative to individuals with no repeat expansion (OR=2.27 [1.82-2.85]).

SNPs at *TMCO1* that tagged expanded VNTR alleles offered the opportunity to replicate these associations in independent, well-powered glaucoma and IOP genetic association data sets (Gharahkhani et al. 2021, Bonnemaijer et al. 2019) (Fig. 3d,e). In these replication cohorts, carriers of rs116089225:C>T, the SNP allele associated with greatest mean VNTR allele length among carriers in UKB (AF=0.01; 11 repeats in a genotyped carrier in HGSVC2; Methods), exhibited significantly elevated glaucoma risk (OR=1.70 [1.43–2.01]; Fig. 3d) and IOP (β=0.201 (0.043) s.d.; Fig. 3e) relative to carriers of the common risk haplotype that segregated with all expanded alleles (AF=0.12, carrier mean allele length = 7.6 repeats in UKB; glaucoma OR=1.34 [1.29–1.39], IOP β=0.084 (0.012) s.d.; Fig. 3d,e). These results from studies that excluded UK Biobank provided confirmatory evidence for the series of VNTR allele effects we observed in UKB.

Though the statistical evidence points to the VNTR as the causal variant driving glaucoma associations at *TMCO1*, the molecular mechanism and causal gene underlying this association remain elusive. Consistent with previous reports (Sharma et al, 2012), carriers of a rare *TMCO1* loss-of-function mutation (rs752176040:ACT>A, AF=0.00034 in exome-sequenced UKB participants; Backman et al. 2021) did not appear to have elevated IOP (β=0.114 (0.135) s.d.) or increased glaucoma risk (OR=1.07 [0.58–1.96]). Analysis of loss-of-function mutation carriers for other nearby genes also did not provide any clues toward a candidate gene (Supplementary Fig. 2). In RNA sequencing data from GTEx (Aguet et al. 2020), VNTR length associated with expression at *TMCO1* in most tissues (e.g., in sun-exposed skin, *P=*2.9 × 10^−11^ for *TMCO1* expression and *P*=8.3 × 10^−26^ for *TMCO1-AS1* expression), consistent with recent SNP-based colocalization analyses (Hamel et al. 2022). However, these associations did not display evidence of an allelic series (Supplementary Fig. 3). Additionally, in joint models including both the VNTR and nearby SNPs, VNTR length did not significantly associate with expression, whereas SNPs retained significance (e.g., *P*=0.5 for the VNTR vs. *P*=0.00011 for rs2790052 for association with *TMCO1-AS1* expression in sun-exposed skin). These results suggest that a variant other than the VNTR, possibly rs2790052 or rs2251768 in the promoter region of *TMCO1*, is responsible for the main eQTL at this locus and that the expression signal is unrelated to the glaucoma and IOP associations.

### Common repeat polymorphism at *EIF3H* associates with a twofold range of colorectal cancer risk

Colorectal cancer is a heritable complex disease for which more than one hundred common risk alleles have been identified, each with a subtle influence on disease risk (OR<1.2) (Huyghe et al. 2019). By contrast, the length of a 27bp repeat (usually ranging from 2-6 repeat units) ∼20kb downstream of *EIF3H* associated strongly with risk of colorectal cancer and colon polyps (*P*= 1.3 × 10^−24^ and *P*=9.3 × 10^−34^, respectively; Fig. 4a,b), with the longest common allele (6 repeat units; AF=0.04) conferring higher colorectal cancer risk (OR=1.34 [1.24–1.45]) than any common SNP or indel in the genome (Fig. 4c). The VNTR appeared to explain nearby SNP associations that were among the first associations reported for colorectal cancer (Tomlinson et al. 2008). Moreover, the explanatory power of this locus, which ranked first among all colorectal cancer loci genome-wide (*P*=1.3 × 10^−24^ for the VNTR vs. *P*=2.2 × 10^−19^ for the strongest SNP association; Fig. 4a), had previously been underestimated by ∼50% in association studies that considered only SNPs which are in partial LD with the VNTR (maximum *R*^2^=0.27; Fig. 4a,b). Imputation of the VNTR association into summary statistics (Pasaniuc et al. 2014) (that excluded UKB) from a large colorectal cancer meta-analysis (Huyghe et al. 2019) replicated the VNTR association as the strongest at the locus (imputed *P*=6.7 × 10^−11^ for the VNTR vs. *P*≥7.3 × 10^−9^ for nearby SNPs; Supplementary Fig. 4). In UKB, the VNTR’s association was driven by a series of four common alleles (3-6 repeat units) which exhibited increasing effects on risk of colorectal cancer and colon polyps. Disease risk increased linearly (on the log-odds scale) with VNTR length (Fig. 4c), with each additional repeat unit associating with a 14% (11–17%) increased risk of colorectal cancer (9% [7–10%] for colon polyps). The effects of an individual’s two alleles appeared to be additive (*P*=0.68 for interaction term), such that common repeat length variation at *EIF3H* appeared to produce a >2-fold range of colorectal cancer risk across individuals (Supplementary Fig. 5).

**Figure 4.**
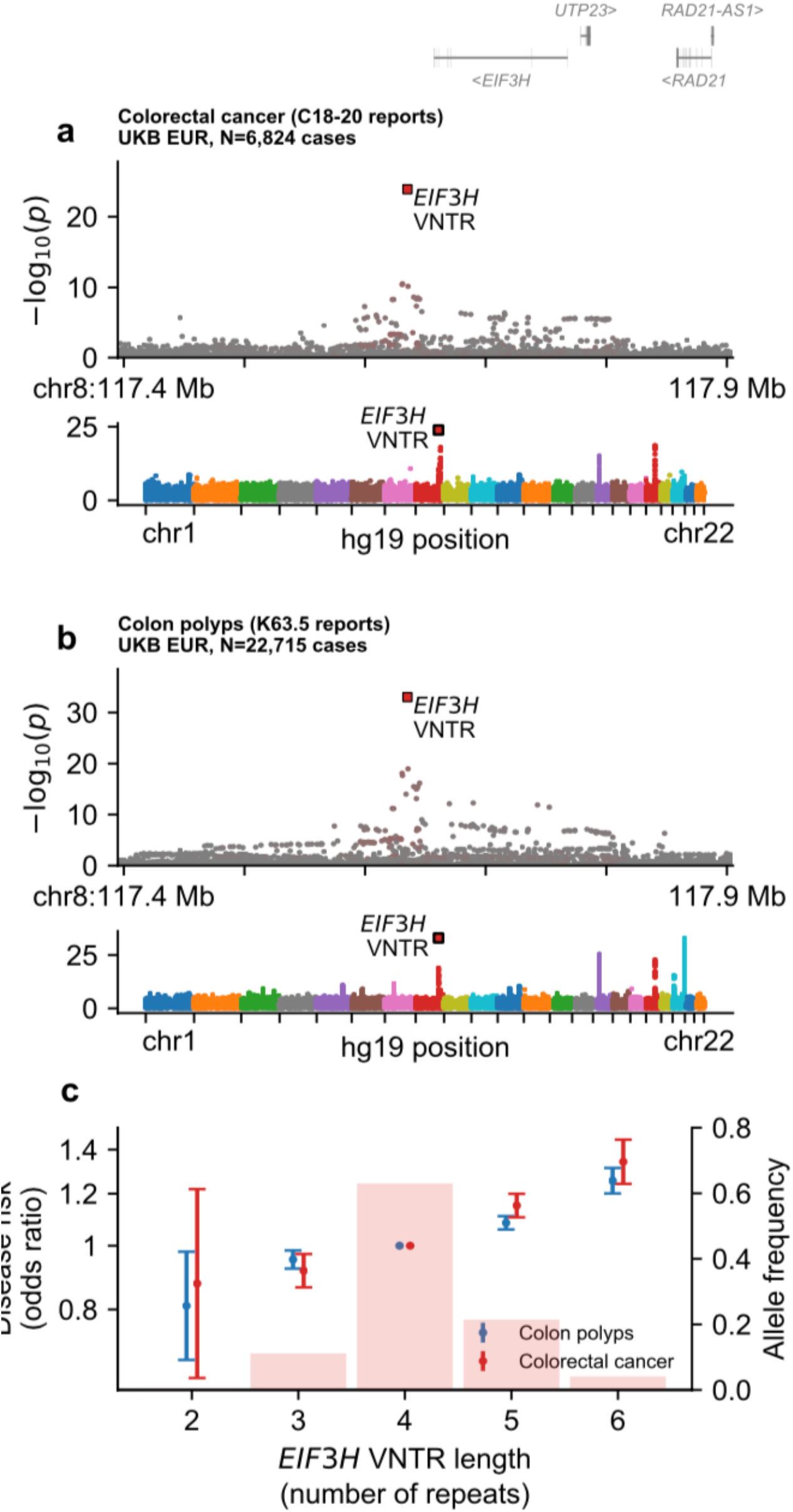
A repeat expansion downstream of *EIF3H* associates with colorectal cancer risk and colon polyps. **a,b)** Associations of inherited variants with colorectal cancer (a) and colon polyps (b) at the *EIF3H* locus (top) and genomewide (bottom). Colored markers in locus plots, variants in partial LD with the VNTR (*R*^2^>0.01). **c)** Frequencies of VNTR alleles observed in European-ancestry UKB participants (histogram, right axis) and their effect sizes (markers, left axis) for colorectal cancer (red) and colon polyps (blue). Error bars, 95% CIs.

The length of this VNTR did not associate with expression of any nearby gene in analyses of RNA sequencing data, either from healthy tissue sequenced by GTEx (Aguet et al. 2020) (*P*≥0.002 for each of 11 genes within 1Mb and each of up to 49 tissues) or from colorectal tumor tissues from the Cancer Genome Atlas (Cancer Genome Atlas Network 2012) (*P*≥0.1 for each of 8 protein-coding genes in analysis of 465 tumor samples). The gene *EIF3H*, which encodes a subunit of a translation initiation factor, has been nominated as a potential causative gene at this locus (Tomlinson et al. 2008). However, definitive evidence linking colorectal cancer risk variants at 8q23.3 to a gene has remained elusive, and, consistent with our findings, risk alleles at this locus have not been shown to associate with *EIF3H* expression (Carvajal-Carmona et al. 2011). Though we have identified the VNTR as a promising candidate for the causal variant at this locus (with statistical support from analyses of two distinct phenotypes – colorectal cancer and colon polyps; between-phenotype *R*^2^=0.02 – as well as independent replication), deciphering the molecular mechanism will require new kinds of data.

### Intronic repeat expansion in *CUL4A* influences alternative splicing and erythrocyte traits

At *CUL4A*, expansion of a highly polymorphic intronic repeat (commonly consisting of ∼3–100 copies of a 29-32bp repeat unit; Fig. 5a) associated with decreased mean corpuscular hemoglobin (*P*=6.4 × 10^−61^, Fig. 5b,c) and nine other erythrocyte-related traits (Fig. 2, Fig. 5c, and Supplementary Table 4). The VNTR association was >3-fold stronger than that of nearby SNPs (none of which could effectively tag the VNTR polymorphism: maximum *R*^2^=0.30; Fig. 5b) and was driven by a series of alleles with monotonically strengthening effects on the associated phenotypes (Fig. 5c). UK Biobank participants carried a multimodal allele distribution with a long tail of expanded alleles (Fig. 5c), consistent with expanded alleles observed in HGSVC2 assemblies (Supplementary Fig. 1). The longest alleles (top 1%, >2.1kb) associated with 0.075 (0.010) s.d. reduced mean corpuscular hemoglobin (Fig. 5c).

**Figure 5.**
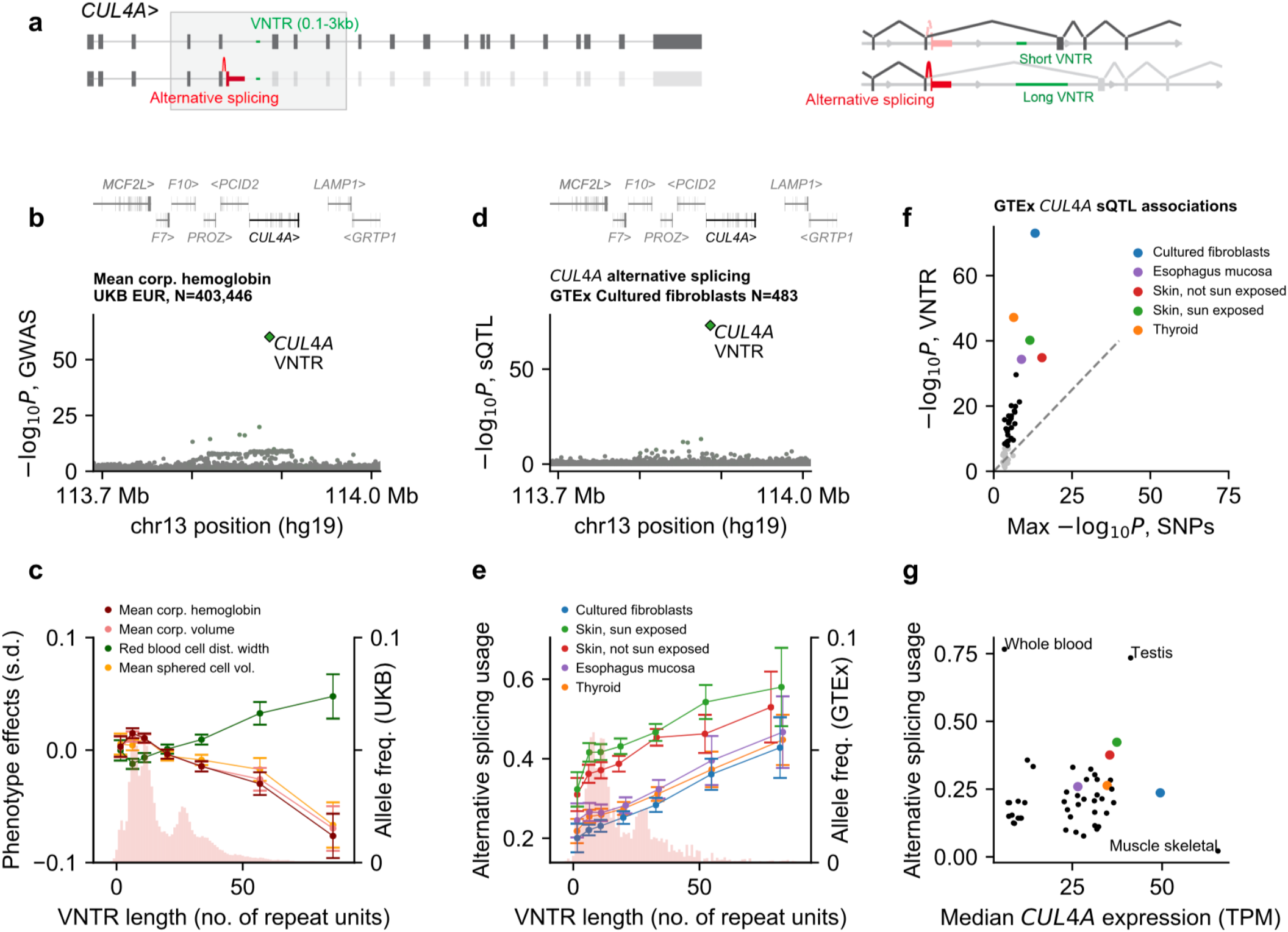
An intronic repeat expansion at *CUL4A* associates with erythrocyte traits and splice isoform usage. **a)** Alternative splicing of two commonly expressed *CUL4A* isoforms. The fifth intron of the canonical transcript contains a highly length-polymorphic VNTR (0.1-3kb, green). The image on the right is zoomed in on the region of *CUL4A* containing the alternative splice. **b)** VNTR and SNP associations with mean corpuscular hemoglobin at the *CUL4A* locus. Colored markers, variants in partial LD with the VNTR (*R*^2^>0.01). **c)** VNTR allele length distribution in European-ancestry UKB participants (histogram, right axis) and mean phenotype in carriers of VNTR alleles (binned by length) for the four most strongly associated blood cell traits (lines, left axis). **d)** VNTR and SNP associations with *CUL4A* alternative splicing usage in cultured fibroblasts. Colored markers, variants in partial LD with the VNTR (*R*^2^>0.01). **e)** VNTR allele distribution in GTEx (histogram, right axis) and mean alternative splice usage in carriers of VNTR alleles (binned by length) for the five tissues with the strongest VNTR association (lines, left axis). Alternative splice usage is the proportion of *CUL4A* transcripts that are alternatively spliced as indicated in panel (a) (as quantified by LeafCutter (Li et al. 2018); Methods). **f)** Scatter plot of VNTR association strength vs. strength of the strongest SNP association with alternative splicing in each of the *N*=49 tissues analyzed by GTEx. Gray dots, tissues for which no variant significantly associated with splicing. **g)** Scatter of median alternative splice usage vs. median *CUL4A* expression for each of *N=*49 tissues. Error bars, 95% CIs.

Analysis of GTEx RNA-seq data revealed that VNTR allele length strongly associated with an apparent splice defect in *CUL4A*, in which individuals carrying longer VNTR alleles were less likely to make the canonical splice over the VNTR in intron 5 (which varies in length from ∼3– 6kb owing to the VNTR polymorphism), instead splicing to a much more proximal sequence (122bp from the splice donor) that results in premature truncation of the *CUL4A* reading frame without the 15 downstream canonical exons (*P*=1.0 × 10^−73^ in cultured fibroblasts; *P≤*0.05 in 47 of 48 additional tissues tested; Fig. 5a,d,e,f). This splice event associated with the VNTR much more strongly than with any SNP, in each of the 30 tissues for which a variant reached Bonferroni significance (Fig. 5f). In each case, longer alleles associated with greater usage of the protein-truncating isoform (Fig. 5e).

The gene *CUL4A* encodes a ubiquitin ligase with an ortholog (*Cul4A*) that is required for hematopoiesis in mice (Waning et al. 2008), suggesting a molecular mechanism in which the VNTR length polymorphism might influence erythrocyte traits by interfering with *CUL4A* splicing and thereby modulating production of a truncated CUL4A isoform with reduced or lost function. Beyond the influence of the VNTR, the proportion of *CUL4A* transcripts that are mis-spliced in this way varies considerably across tissues (from an average of ∼0.03 in skeletal muscle to ∼0.75 in whole blood and testis; Fig. 5g), suggesting that cellular context, as well as VNTR length, affect splicing outcome.

### Non-coding repeat polymorphisms near *SIRPA, DOCK8*, and *PLEC* associate with platelet traits

The three strongest non-coding VNTR associations supported by statistical fine-mapping involved repeats near *SIRPA, DOCK8* and *PLEC*, with each VNTR appearing to underlie one of the top 40 associations genome-wide for a platelet phenotype, explaining >0.1% of variance (Fig. 2, Supplementary Fig. 6, and Supplementary Table 4). Approximately 50kb upstream of *SIRPA*, a 45bp repeat (with common 6-repeat and 10-repeat alleles) residing within a predicted enhancer of *SIRPA* (Fishilevich et al. 2017) associated with mean platelet volume (MPV; *P*=1.6 × 10^−167^; FINEMAP PIP=1.00). Within *DOCK8*, a gene harboring rare coding variants previously linked to MPV (Barton et al. 2021), the length of a highly length-polymorphic (∼100bp–6kb) intronic 61bp repeat also associated strongly with MPV (*P*=6.5 × 10^−129^; PIP=1.00). At *PLEC*, which encodes plectin, an intermediate filament binding protein with roles in cytoarchitecture and cell shape (Svitkina et al. 1996), the length of a 76bp intronic repeat (2-13 repeats per allele) associated with platelet distribution width (*P*=1.2 × 10^−96^; PIP=0.99).

Other fine-mapped associations provided additional examples of ways in which previously-hidden repeat polymorphisms appear to influence human phenotypes. VNTR length polymorphisms in introns of *CHMP1A* and *SBNO2* associated with decreased hypertension risk (*P*=1.5 × 10^−12^; PIP=1.00) and bone mineral density (*P*=3.4 × 10^−45^; PIP=0.97), respectively, apparently by modulating splicing of these genes (Supplementary Figs. 7 and 8). Bone mineral density was also significantly associated (*P*=1.8 × 10^−12^; PIP=1.00; Supplementary Fig. 6) with length of a VNTR in the promoter region of *ITGB2* (encoding CD18), a gene which previous work has implicated in osteogenesis (Miura et al. 2005). These associations provide plausible bases for further investigation of the function of tandem repeat polymorphisms in specific clinical contexts.

## Discussion

These results identify many VNTRs that appear to have strong effects on human phenotypes, including five VNTR length polymorphisms that are associated with risk of common diseases. Two disease associations we observed, involving VNTRs at *TMCO1* and *EIF3H*, appeared to be the strongest known genetic influences of common inherited variation on glaucoma and colorectal cancer risk, respectively. These discoveries were enabled by a computational approach to VNTR genotype estimation that integrated sequencing depth-of-coverage analysis with statistical phasing and imputation into SNP-array genotyping data – a framework that can similarly be applied to other genetic data sets.

Most of the phenotype-associated VNTRs identified by our study lie in non-coding regions of the genome, modifying DNA sequence length by hundreds to thousands of base pairs, yet with no obvious molecular mechanism to explain their apparent impact on phenotype. A notable exception is the hemoglobin-associated VNTR we identified in *CUL4A*, which appeared to strongly attenuate the excision of the intron in which it resides, drastically affecting the mature mRNA sequence and, presumably, the function of the translated protein. Non-coding variants linked to phenotype pose a central challenge in human genetics. Consistent with other studies, available RNA sequencing data sets helped interpret only a small fraction of the non-coding associations we observed. Other techniques and further study will be required to elucidate the “missing regulation” and identify the mechanisms underlying the associations observed here (Connally et al. 2021).

Despite considerably expanding the set of known associations between VNTRs and human phenotypes, the results presented here likely represent an incomplete look into the landscape of repeat-mediated trait heritability, owing to genotyping challenges that we could only partially overcome as well as inherent limitations of the UK Biobank cohort we analyzed. Our read-depth and haplotype-modeling approach accurately captured larger-scale VNTR length variation (>100bp) but produced noisier genotype estimates for less-variable VNTRs, reducing power to analyze such VNTRs. Moreover, we excluded all short tandem repeat (STR) loci from analysis, for which other methods are required (Saini et al. 2018, Margoliash et al. 2022). The set of VNTRs we considered was also limited by our GRCh38-based VNTR ascertainment strategy (which required multiple repeat units to be present in the human reference) and the need for mappability of short reads. Additionally, our ability to detect VNTR-phenotype associations was limited by the demographics of the UK Biobank cohort, which enrolled generally healthy participants of predominantly European ancestry. Analyses in case-control cohorts enriched for heritable diseases will be needed to power discovery of further influences of VNTR variation on human health. Many of these limitations are now beginning to be overcome as long-read sequencing data sets scale to thousands of samples (Beyter et al. 2021) and short-read WGS data sets scale to hundreds of thousands of samples (Halldorsson et al. 2022), including in diverse populations (All of Us Research Program 2019). We anticipate that these recently-generated and upcoming data resources will enable many further insights into the contribution of repeat polymorphisms to heritable complex traits in the years to come.

## Supporting information

Supplementary Notes (with references)

## Data Availability

Individual-level VNTR genotypes imputed into UKB will be returned to the UK Biobank Resource. The VNTR+SNP reference panel in SSC will be returned to SFARI Base. Summary statistics for VNTR-phenotype association tests are available at https://data.broadinstitute.org/lohlab/UKB_genomewideVNTR_sumstats/. Access to the following data resources used in this study is available to all approved researchers upon application: UK Biobank (http://www.ukbiobank.ac.uk/), Simons Simplex Collection (SSC Whole- genome 2, https://base.sfari.org), TCGA (via dbGaP, https://www.ncbi.nlm.nih.gov/gap/, accession phs000178.v11.p8), GTEx (via dbGaP, https://www.ncbi.nlm.nih.gov/gap/, accession phs000424.v8.p2; the GTEx Portal http://www.gtexportal.org). The following data resources used in this study are publicly available: 1000 Genomes Project (including HGSVC2 long-read assemblies, http://www.internationalgenome.org/), NHGRI-EBI GWAS Catalog (http://ebi.ac.uk/gwas/, accessions GCST009413, GCST90011767, and GCST012879).

## Acknowledgments

We would like to thank A. Segre, A. Lee, N. Kamitaki, and R. Gupta for helpful discussions related to this work. This research was conducted using the UK Biobank Resource under application #40709. Computational analyses were performed on the O2 High Performance Compute Cluster, supported by the Research Computing Group, at Harvard Medical School (http://rc.hms.harvard.edu) and the UK Biobank Research Analysis Platform (RAP). We are grateful to all of the families at the participating Simons Simplex Collection (SSC) sites, as well as the principal investigators (A. Beaudet, R. Bernier, J. Constantino, E. Cook, E. Fombonne, D. Geschwind, R. Goin-Kochel, E. Hanson, D. Grice, A. Klin, D. Ledbetter, C. Lord, C. Martin, D. Martin, R. Maxim, J. Miles, O. Ousley, K. Pelphrey, B. Peterson, J. Piggot, C. Saulnier, M. State, W. Stone, J. Sutcliffe, C. Walsh, Z. Warren, E. Wijsman). We appreciate obtaining access to genetic data on SFARI Base. The results presented here are in part based upon data generated by the TCGA Research Network: https://www.cancer.gov/tcga. The Genotype-Tissue Expression (GTEx) Project was supported by the Common Fund of the Office of the Director of the National Institutes of Health, and by NCI, NHGRI, NHLBI, NIDA, NIMH, and NINDS.

## Author contributions

R.E.M., R.E.H., S.A.M., and P.-R.L. conceived and designed the study. R.E.M., R.E.H., and P.-R.L. designed and implemented the statistical methods and performed the computational analyses. R.E.M., R.E.H, A.R.B., M.A.S., M. L. A. H., S.A.M., and P.-R.L interpreted analytical results. All authors wrote and edited the manuscript.

## Data availability

Individual-level VNTR genotypes imputed into UKB will be returned to the UK Biobank Resource. The VNTR+SNP reference panel in SSC will be returned to SFARI Base. Summary statistics for VNTR-phenotype association tests are available at https://data.broadinstitute.org/lohlab/UKB_genomewideVNTR_sumstats/. Access to the following data resources used in this study is available to all approved researchers upon application: UK Biobank (http://www.ukbiobank.ac.uk/), Simons Simplex Collection (SSC Whole-genome 2, https://base.sfari.org), TCGA (via dbGaP, https://www.ncbi.nlm.nih.gov/gap/, accession phs000178.v11.p8), GTEx (via dbGaP, https://www.ncbi.nlm.nih.gov/gap/, accession phs000424.v8.p2; the GTEx Portal http://www.gtexportal.org). The following data resources used in this study are publicly available: 1000 Genomes Project (including HGSVC2 long-read assemblies, http://www.internationalgenome.org/), NHGRI-EBI GWAS Catalog (http://ebi.ac.uk/gwas/, accessions GCST009413, GCST90011767, and GCST012879).

## Code availability

The following publicly available software resources were used to perform analyses in this work BOLT-LMM (v2.3.6), https://data.broadinstitute.org/alkesgroup/BOLT-LMM/; FINEMAP (v1.3.1), http://www.christianbenner.com/; plink (v1.9 and v2.0), https://www.cog-genomics.org/plink2/; Tandem Repeats Finder (v4.09.1), tandem.bu.edu/trf/trf.html; minimap2 (v2.18-r1015), https://github.com/lh3/minimap2; fastQTL (v2.0), https://github.com/francois-a/fastqtl/; Genome STRiP, https://software.broadinstitute.org/software/genomestrip/.

## Methods

### UK Biobank genetic data

The UK Biobank resource contains extensive genetic and phenotypic data for ∼500,000 participants recruited from across the UK (Sudlow et al. 2015). We analyzed SNP and indel genotypes available from blood-derived SNP-array genotyping of 805,426 variants in 488,377 participants and subsequent imputation to 93,095,623 autosomal variants (using the Haplotype Reference Consortium and UK10K + 1000 Genomes Phase 3 reference panels) in a subset of 487,409 participants (Bycroft et al. 2018). We further analyzed alignments from whole-genome sequencing (WGS) of 200,018 participants (>20x coverage by 151bp paired-end reads) (Halldorsson et al. 2022).

### UK Biobank phenotype data

We performed initial analyses on a set of 786 phenotypes (Supplementary Table 2) that we curated from the UK Biobank “core” data set as described in (Mukamel et al. 2021). This set of phenotypes consisted of: (i) 636 diseases collated by UKB from several sources (self-report and accruing linked records from primary care, hospitalizations, and death registries) into single “first occurrence” data fields indexed by ICD-10 diagnosis codes; and (ii) 150 continuous and categorical traits selected based on high heritability or common inclusion in genome-wide association studies. Phenotypes in the latter set were derived from physical measurements and touchscreen interviews; blood count, lipid and biomarker panels of biological samples; and follow-up online questionnaires. For continuous traits, we performed quality control and normalization (outlier removal, covariate adjustment, and inverse normal transformation) as previously described (Barton et al. 2021, Loh et al. 2018). In follow-up analyses at *TMCO1* and *EIF3H*, we refined associated disease phenotypes (related to ICD-10 codes H40 and K63) and curated related phenotypes (intraocular pressure and colorectal cancer) not in our initial analysis set (Supplementary Note).

### Simons Simplex Collection genetic data

We analyzed aligned sequencing reads and the hg38 variant call set derived from SSC WGS 2. The SSC cohort consists of individuals from 2,600 families, each of which has one child affected with an autism spectrum disorder. Each participant was deeply whole-genome sequenced (30x mean coverage, 150bp paired-end reads). We analyzed genetic data obtained from a subset of 8,936 participants, which included 1,901 quartets (parents, proband, and unaffected sibling) and 440 trios (parents and child). In total, the analysis set contained 4,688 unrelated parents whose 9,376 haplotypes we included in our VNTR+SNP reference panel. We applied multiple rounds of quality control to generate a high-quality set of phased SNP haplotypes for SSC participants (Supplementary Note).

### Sample filters for ancestry and relatedness

For genetic association analyses in UKB, we applied strict filters to avoid confounding from population stratification and relatedness among individuals. We performed all analyses on a filtered set of 418,136 individuals that we identified by: (i) removing principal component (PC) outliers (more than six standard deviations from the mean among individuals who reported White ethnicity in any of the first 10 genetic PCs); and (ii) removing one individual from each ≤2^nd^-degree related pair (kinship coefficient > 0.0884) previously identified by UKB (Bycroft et al. 2018).

For benchmarking accuracy of VNTR length estimation in SSC, we assigned ancestry to SSC participants using the software SNPweights v2.1 (Chen et al. 2013). Using pre-computed SNP weights for European, West African, East Asian and Native American ancestral populations (accessed from https://cdn1.sph.harvard.edu/wp-content/uploads/sites/181/2014/03/snpwt.NA_.zip on 08/20/2019), we estimated the proportion of each SSC participant’s genome that derived from each ancestral population. We identified 3,904 unrelated parents whose genetic ancestry was estimated to be largely (>80%) European.

### VNTR ascertainment and genotyping pipeline

We identified VNTR loci and genotyped VNTR allele length variation from whole-genome sequencing data using an analysis pipeline consisting of three main steps (detailed in the Supplementary Note):

1. ***Identify VNTR loci from analysis of the human reference and HGSVC2 long-read assemblies***. We started by searching the GRCh38 reference for tandem repeats using two approaches:
  a. Tandem Repeats Finder (Benson 1999) v4.09, which we ran using its suggested parameters 2 5 7 80 10 50 2000 -l 6 -h to detect patterns up to 2kb. We filtered to autosomal repeats with length ≥100bp, period ≥10bp, #repeats ≥2, and sequence identity ≥75%, and we applied a rough deduping of duplicated regions (overlap (intersection/union) >75% or both endpoints within 75bp); and
  b. VNTRScanner and VNTRPartitioner, algorithms we had previously developed to detect larger repeats with potentially greater variability within the repeat units (Mukamel et al. 2021).
  The combination of the two methods resulted in an initial set of 100,844 autosomal repeat loci with an estimated repeat unit ≥7bp long. We then analyzed each tandem repeat in 64 HGSVC2 long-read-based haploid genome assemblies (Ebert et al. 2021) to identify which regions were multiallelic. We filtered to loci with ≥3 distinct alleles represented among the 64 HGSVC2 long-read assemblies, applied several additional quality control filters, and removed duplicated regions (using benchmarks from SSC WGS to adjudicate among substantially-overlapping regions), resulting in 15,653 VNTR loci for further analysis (Supplementary Note).
2. ***Estimate VNTR lengths from WGS depth-of-coverage in SSC***. At each VNTR, we estimated diploid VNTR content (i.e., the sum of VNTR lengths across an individual’s two alleles) for SSC participants by analyzing the aligned WGS reads overlapping the VNTR using Genome STRiP (Handsaker et al. 2015), using dosage estimates from normalized read depth to estimate VNTR length (summed across parental alleles). We corrected for observed batch effects using Leiden clustering. We benchmarked the resulting (pre-refinement) VNTR genotypes using measurements from related individuals in SSC, and applied several additional variant-level quality control filters derived from these benchmarks (Supplementary Note).
3. ***Phase and impute VNTR allele length estimates by modeling haplotype sharing***. We performed statistical phasing on estimates of diploid VNTR content to estimate haploid allele lengths, and we used the resulting VNTR+SNP haplotypes for imputation of VNTR allele lengths into the UKB cohort. To do so, we adapted the computational algorithm we had previously used (Mukamel et al. 2021; Supplementary Note).

Of the 15,653 multiallelic VNTR loci identified in the first step of this pipeline, we identified a subset of 9,561 loci suitable for downstream association analyses in UKB based on genotyping quality (estimated *R*^2^>0.1), imputation quality (estimated *R*^2^>0.1), and other quality control filters (Supplementary Note; Supplementary Table 1).

We also considered applying other methods previously developed for genotyping repeats such as adVNTR (Bakhtiari et al. 2018) and ExpansionHunter (Dolzhenko et al. 2017). However, we were unable to use adVNTR because it requires sequencing reads to span a VNTR, and the majority of VNTR loci (90%) had an allele longer than the read length in SSC (150bp). While ExpansionHunter is capable of genotyping repeats longer than the read length, it was designed primarily for STR genotyping and assumes that different repeat units are mostly identical in sequence, whereas many VNTRs exhibit repeat motif variability (Course et al. 2021). Beyond needing to overcome these specific limitations, we were also motivated to apply methods that leverage haplotype sharing among unrelated individuals in large cohorts to refine genotypes, increasing power to detect downstream associations (Mukamel et al. 2021).

### VNTR-phenotype association and fine-mapping analyses

We performed association tests between the 9,561 imputed VNTRs and 786 phenotypes in our analysis set of 418,136 unrelated UK Biobank participants of genetically-determined European ancestry (see above). We computed linear regression association statistics using BOLT-LMM (Loh et al. 2015) v2.3.6 including a standard set of covariates (20 genetic PCs, assessment center, genotyping array, sex, age, and age^2^), and found 4,968 VNTR-phenotype pairs that passed a Bonferroni-corrected significance threshold of *P*<5 × 10^−9^ (reflecting the ∼10,000 VNTRs x ∼1,000 phenotypes we tested for association; Supplementary Table 3). Linear regression produced well-calibrated *P*-values given that the VNTRs we analyzed exhibited common multiallelic variation and the binary phenotypes we analyzed were not ultra-rare (at least a few hundred cases in UKB; Mukamel et al. 2021).

To determine which of these VNTR-phenotype associations were likely to represent causal effects of VNTR allele length variation (vs. tagging of nearby causal SNPs), we first computed linear regression association statistics for all nearby SNPs and indels imputed by UKB (within 500kb of the VNTR) as we had for the VNTR. We then applied the Bayesian fine-mapping software FINEMAP (Benner et al. 2016) v1.3.1 (options -- corr-config 0.999 --sss --n-causal-snps 5) to estimate the likelihood of causality for the VNTR, accounting for linkage disequilibrium with 500 of the most strongly associated nearby variants. For fine-mapping, we excluded SNPs at multiallelic sites, rare variants (MAF<0.001), and variants called within the VNTR region. For associations for which the VNTR was assigned a high posterior probability of causality (PIP>0.5), we ran a second round of fine-mapping including 2,000 of the most strongly associated nearby variants. The results of these analyses are summarized in Supplementary Table 3. In total, 107 VNTR-phenotype associations involving 58 distinct VNTRs were assigned a high posterior probability of causality by FINEMAP (PIP>0.5 in both rounds; Supplementary Table 4, Fig. 2).

We additionally tested each VNTR for association with autism directly in SSC. To do so, we computed linear regression association statistics, restricting to the probands and siblings in 1,901 complete quartets and including sex as a covariate. None of the VNTRs we tested reached our study-wide significance threshold (*P*<5 × 10^−9^).

For VNTRs at loci of particular interest, we ran additional analyses after improved genotyping and refined phenotyping of disease traits as detailed in the Supplementary Note.

### SNP-based corroboration of TMCO1 VNTR length associations with glaucoma and intraocular pressure in independent cohorts

Previous genome-wide association studies of glaucoma and intraocular pressure provided the opportunity to replicate (in part) the allelic series we observed at *TMCO1* in UK Biobank (in which *TMCO1* VNTR alleles of increasing length associated with increasing glaucoma risk and IOP). To do so, we identified SNPs that tagged VNTR alleles of different lengths and then examined their effect sizes in publicly available summary association statistics from the NHGRI-EBI GWAS Catalog (Buniello et al. 2019) for study GCST90011767 (Gharahkhani et al. 2021; glaucoma GWAS, downloaded on 07/15/2022) and GCST009413 (Bonnemaijer et al. 2019; IOP GWAS, downloaded on 07/21/2022). These studies did not include UK Biobank data and were thus suitable for independent replication.

For this corroboratory analysis, we sought to identify a SNP that segregated with particularly long VNTR alleles and then compare its effect size to that of rs2790053, a representative of the common risk haplotype (AF=0.12) that segregates with all expanded alleles (5 or more repeats; Fig. 1a) – the idea being that a SNP tagging extra-long VNTR alleles should associate with even higher glaucoma risk and IOP than the common risk haplotype that tags a mixture of all long alleles.

To find such a SNP, we examined heterozygous carriers of each SNP within 500kb of the VNTR with MAF between 0.1% and 10% and computed the mean length of the longer allele present in each carrier (which should almost always be the desired allele for SNPs that tag very long VNTR alleles). We performed this analysis using imputed genotypes available for UK Biobank participants of European ancestry (from the UKB imp_v3 data set (Bycroft et al., 2018), reasoning that summary statistics available from previous GWAS rely on similar imputation. We restricted analysis to individuals in the UKB *N*=200K WGS data set and used allele lengths that we estimated via our optimized genotyping of *TMCO1* (Supplementary Note).

Upon ranking SNPs in descending order of mean estimated VNTR length in carriers, two low-frequency SNPs – rs35310077 and rs116089225, the top two SNPs on the list – were clearly the best candidates for replication, segregating with VNTR alleles with mean estimated lengths of ∼9.7 repeat units. (This length estimate is probably downward-biased by imputation error resulting in regression to the mean; a single carrier of both SNPs in HGSVC2 carried an 11-repeat allele.) Closer inspection of these two SNPs showed that rs35310077 (MAF=0.007 in UKB imp_v3) tagged a sub-haplotype of rs116089225 (MAF=0.009 in UKB imp_v3), with the latter SNP exhibiting higher imputation accuracy (INFO=0.95 for rs116089225 vs. INFO=0.89 for rs35310077). We therefore proceeded with rs116089225 (and rs2790053, the aforementioned tag SNP for the AF=0.12 common risk haplotype) for lookup in glaucoma and IOP summary statistics.

### Imputation-based corroboration of EIF3H VNTR length association with colorectal cancer risk

We sought to replicate the association we observed between *EIF3H* VNTR length and colorectal cancer risk by imputing the VNTR’s association statistic into SNP association statistics from an independent colorectal cancer GWAS. We employed the approach of ImpG (Pasaniuc et al. 2014), which estimates a variant’s association statistic based on the association statistics of variants in LD (using a multivariate normal with covariance derived from the LD matrix). Summary statistics were downloaded from the NHGRI-EBI GWAS Catalog (Buniello et al. 2019) for study GCST012879 (Huyghe et al. 2019, accessed on 07/15/2022). We extracted statistics for all SNPs within 300kb of the VNTR that were also present in the UKB imp_v3 data set (Bycroft et al. 2018). We computed LD using VNTR+SNP genotypes for 16,728 UKB participants, obtained by 25x-downsampling the set of 418,136 individuals used in our association analyses. *EIF3H* VNTR genotypes were estimated from refined genotyping using UKB *N=*200K WGS data (Supplementary Note). Since the published implementation of ImpG requires variants to be biallelic, we re-implemented the method (using the same default regularization parameter L=0.1) to apply it to continuous-valued VNTR allele length estimates; we previously validated this re-implementation of ImpG (Mukamel et al. 2021). Consistent with our observations in UK Biobank, the imputed association statistic for the VNTR in these independent summary statistics (from Huyghe et al. 2019, excluding UKB) was larger than that of any nearby SNP or indel (imputed *P*=6.7 × 10^−11^ for the VNTR vs. *P*=7.3 × 10^−9^ for to the top SNP at the *EIF3H* locus).

### Expression and splicing quantitative trait association analyses in GTEx

We performed expression and splicing quantitative trait association analyses using data from the Genotype-Tissue Expression (GTEx) project (V8). The GTEx project analyzed 49 human tissues, measuring DNA and RNA collected from 15,201 biosamples contributed by 838 post-mortem donors (Aguet et al. 2020). We estimated VNTR allele lengths for GTEx participants by imputation into WGS-derived SNP genotypes previously phased by GTEx using SHAPEIT2 (Delaneau et al. 2013) (accessed on 07/22/2021 via the Terra data platform) and, at certain loci, WGS read alignments.

Specifically:

- At *EIF3H, CHMP1A* and *SBNO2*, we imputed VNTR allele lengths using the reference VNTR+SNP haplotypes and imputation parameters we obtained from analysis of SSC.
- At *TMCO1*, we adapted the strategy for improved genotyping that we used in UKB, combining read-level information from WGS reads spanning short alleles, counts of WGS reads internal to long alleles, and nearby SNP genotypes (Supplementary Note).
- At *CUL4A*, we imputed VNTR allele lengths using reference VNTR+SNP haplotypes and imputation parameters we obtained from analysis of *N*=200K WGS UKB samples (Supplementary Note). We imputed into SNP haplotypes that we rephased using Eagle2 (Loh et al. 2016) (--Kpbwt=100000, -- pbwtIters=3) using the full UKB cohort (*N*∼487K) as a reference panel, restricting analysis to variants typed on the UKB SNP-array with concordant EUR allele frequencies (absolute difference <0.1). For analyses at *CUL4A* that did not require haplotype-resolved estimates (Fig. 5d,f), we estimated the diploid content of the highly polymorphic VNTR directly from depth-of-coverage of aligned whole-genome sequencing reads (Supplementary Note), providing estimates which sibling-derived benchmarks in SSC indicated were more accurate than imputed values.

To quantify association strengths with expression and splicing quantitative traits, we emulated the analyses performed by GTEx (Aguet et al. 2020): we obtained normalized expression and splicing quantitative trait phenotypes, as well as covariates, from the GTEx Portal (https://gtexportal.org/home/datasets (V8) accessed on 08/01/2021 and 09/08/2021), and used the software fastQTL v2.0 (Ongen et al. 2016) to compute linear regression VNTR association statistics. We quantified (unnormalized) alternative splice usage at *CUL4A* (Fig. 5e,g) using the intron excision ratio

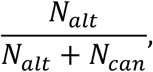

where *N*_*alt*_ and *N*_*can*_ are counts of reads from LeafCutter (Li et al. 2018) (accessed on 12/12/2021 from Terra) for reads supporting excision of the alternative intron (chr13:113229519-113229642) and the canonical intron (chr13:113229519-113233177), respectively (Fig. 5a). We obtained tissue-level estimates of median *CUL4A* expression (Fig. 5g) from the GTEx Portal (accessed from https://storage.googleapis.com/gtex_analysis_v8/rna_seq_data/GTEx_Analysis_2017-06-05_v8_RNASeQCv1.1.9_gene_median_tpm.gct.gz on 07/08/2022).

### Expression quantitative trait association analyses in TCGA

We performed expression quantitative trait association analysis in colorectal cancer tissue using data from the Cancer Genome Atlas (TCGA; Cancer Genome Atlas Network 2012). We obtained phased SNP data from the National Cancer Institute’s Genomic Data Commons (GDC, http://gdc.cancer.gov/, accessed on 03/31/2022), previously generated (Sayaman et al. 2021) using the TOPMed imputation server. For VNTR imputation, we used VNTR+SNP reference haplotypes and parameters obtained from analysis of *EIF3H* in UKB *N=*200K WGS samples (Supplementary Note). We obtained gene expression quantification, measured in fragments per kilobase per million mapped reads (FPKM), in colorectal cancer tissue derived from RNA-sequencing via the GDC portal (http://portal.gdc.cancer.gov/, accessed on 08/23/2022; files matching search terms primary site=colon or rectum, program=TCGA, sample type=primary tumor, workflow=STAR – Counts, data category=transcriptome profiling, data format=tsv, and data type=gene expression quantification). We performed linear regression association tests to quantify the strength of association between imputed VNTR lengths and expression at each of 8 measured genes within 1Mb of the VNTR (465 samples with imputed VNTR lengths and expression data available).

### Assessing the impact of LOF variants near TMCO1 on glaucoma and IOP

We identified variants predicted to cause loss-of-function (pLoF) of genes near *TMCO1* using data derived from whole-exome sequencing of 454,787 UK Biobank participants (Backman et al. 2021). We first extracted a set of pLoF variants for each gene, selecting variants previously annotated as “LoF” by SnpEff. We then used plink2 (Chang et al. 2015) to extract carriers of each pLoF variant from the 450k interim release of population level exome OQFE variants derived from WES. We computed effects on glaucoma risk via logistic regression (including age, age^2^, sex and 20 PCs as covariates) and effects on IOP by taking the phenotypic mean among carriers (adjusted for the same covariates) (Supplementary Fig. 2).

