## Supplementary Notes (with references) for "Repeat polymorphisms in non-coding DNA underlie top genetic risk loci for glaucoma and colorectal cancer"

### 1 Quality-control, phasing, and IBD-calling in SNP data from SSC WGS

We applied multiple rounds of QC to the SSC WGS 2 hg38 variant call set (9,209 samples; An et al. 2018) to facilitate generating a high-quality set of phased SNP-haplotypes for SSC participants.

We first applied several variant-level filters:

- Restricted to biallelic SNPs with MAC>=5 and missingness <0.05.
- Excluded SNPs with allele frequencies (in European-ancestry SSC participants; see Methods) that differed by >0.1 compared to allele frequencies in the UK10K+1000G reference panel (Huang et al. 2015) (subsetting 1000G samples to EUR and lifted from hg19 to hg38) or to allele frequencies in the UK Biobank SNP-array data set (restricted to the British-ancestry subset curated by UK Biobank (Bycroft et al. 2018) and lifted from hg19 to hg38).
- Excluded SNPs with 10 or more Mendelian errors among parent-child trios (computed using the bcftools +mendelian plugin).

We then phased the filtered SNPs using Eagle2 (Loh et al. 2016) and post-processed the phased haplotypes to incorporate trio relationships using the bcftools +trio-phase plugin (a component of the MoChA software package).

We subsequently observed that the set of SNPs that passed the above filters still included a small fraction of SNPs that appeared to have high error rates (often having high rates of heterozygous genotype calls and/or clustered in regions of the genome that did not lift between hg19 and hg38 and thus were not considered in the allele frequency check).

To detect remaining bad SNPs, we therefore implemented an additional round of QC consisting of a Hardy-Weinberg equilibrium check (filtering SNPs with z-score > 5 for observed – expected heterozygotes) and a haploid Mendelian error check (filtering SNPs with >10 disagreements between phased haplotypes transmitted from parents to children in parent-child trios in the SSC data set). To facilitate the latter check, we implemented a simple hidden Markov model (HMM) to match computationally phased haplotypes of each child to computationally phased haplotypes of the child’s two parents (with states corresponding to haplotype assignments, transitions modeling phase switch errors or recombinations, and emissions modeling genotype errors (treated as 10-fold less costly than state changes)). This algorithm allowed us to tabulate the number of haploid Mendelian errors observed at each SNP (based on the Viterbi decoding of each child’s HMM).

After applying the additional two filters above (which together excluded ~1% of the SNPs that had passed the previous QC filters), we then reran phasing using Eagle2 followed by bcftools +trio-phase (in two rounds, first using one sibling from each quartet and then using the remaining sibling from each quartet, which appeared to improve performance). Finally, we reran the HMM above to obtain an “IBD map” matching phased haplotype segments of each child to haplotypes of the child’s two parents for use in downstream analyses.

##### *Sample exclusions for downstream analysis*

Although the SSC WGS 2 variant call set contained 9,209 individuals, only 9,100 of these individuals had accessible sequence alignments (i.e., cram files) needed for downstream WGS read-depth analysis. Among these 9,100 individuals, we excluded 160 individuals whose whole-genome sequencing had been performed in a pilot WGS analysis with read-depth characteristics very different from the remainder of the data set. We further excluded 4 individuals who withdrew from SSC, leaving 8,936 individuals (including 1901 full quartets) for downstream analysis.

#### 2 Estimating diploid VNTR content from WGS read depth in SSC

For each of 100,844 repeat loci we ascertained from the GRCh38 reference (Methods), we estimated diploid VNTR content (i.e., the sum of VNTR lengths across an individual's two alleles) for SSC participants by analyzing the aligned WGS reads overlapping the VNTR using Genome STRiP (Handsaker et al. 2015). We estimated diploid VNTR content using dosage estimates from normalized read depth, without running the Genome STRiP Gaussian mixture model to determine integer copy number.

We benchmarked these VNTR content estimates by analyzing the results from siblings in SSC. We computed the following QC metrics:

1. **IBD2R**. The Pearson correlation between VNTR content measurements of SSC siblings that are identical-by-descent (IBD2) at a given VNTR locus. This quantity was used to estimate the amount of genetic signal that could be ascertained from read-depth analysis at each VNTR.
2. **PARENTR**. The correlation between VNTR content measurements among parents of IBD2 siblings (which was an indicator of batch effects, as this quantity should otherwise be close to zero).
3. **RDIFF**. The difference between IBD2R and PARENTR, i.e., RDIFF=IBD2R-PARENTR. RDIFF is meant to capture the degree to which correlation between sibling measurements captures true genetic variation, rather than technical artifacts (e.g., batch effects) from sequencing.
4. **FLANKR**. For each VNTR locus, we measured read depth within two 1kb segments on each side of the VNTR locus, each separated by 100bp from the VNTR. We then computed the maximum of the Pearson correlation between the read-depth measurements of the segments outside of the VNTR to the read-depth measurement of the VNTR itself. This was used to control for VNTRs occurring within larger copy number variable regions.

We observed that diploid VNTR content estimates in SSC appeared to have significant batch effects, which were only partially correlated with SSC sequencing wave. To better control for these batch effects, we first excluded 160 SSC participants whose whole-genome sequencing had been performed in a pilot WGS analysis with read-depth characteristics very different from the remainder of the data set. To correct for additional batch effects, we then clustered the remaining samples as follows:

1. We selected 2,655 VNTR loci across the genome that (a) had strong evidence of batch effects (PARENTR > 0.1) and (b) were unlikely to be truly polymorphic (absolute value of RDIFF < 0.1).

2. We computed the top 100 principal components (PCs) based on read-depth estimates at these loci, and performed k-nearest-neighbor (k=25) clustering on the samples based on their coordinates in PC-space.
3. We performed Leiden clustering with resolution parameter R=1.0 on the neighbor graph, which partitioned the SSC samples into 18 clusters.

We normalized VNTR content estimates by scaling the estimates within each cluster so the median content across clusters was equal to the population median length. This clustering method and parameters described above were selected after evaluation of several strategies (hclust2 and cutree functions in R, and Louvain and Leiden clustering). We found that Leiden clustering produced the greatest decrease in PARENTR (indicating successful correction of batch effects).

#### 3 Identifying VNTR loci by analysis of human reference and HGSVC2 long-read assemblies

We analyzed the 100,844 repeat regions we identified in GRCh38 (Methods) in HGSVC2 long-read haploid genome assemblies (Ebert et al. 2021) to determine which were multiallelic VNTR loci. As a preliminary step, we removed duplicate loci with greater than 50% reciprocal overlap, prioritizing the loci to keep based on IBD2 sibling correlation in SSC (see Section 2 above). For each remaining repeat locus, we attempted to measure repeat length in each assembly by mapping surrounding sequence from GRCh38 to the assembly.

In detail, we extracted the flanking sequence from the reference (1kb upstream and downstream) using bedtools v2.27.1 (Quinlan and Hall 2010) and aligned the flanks to the assembly using minimap2 v2.18-r1015 (Li 2018) (options --cs -x map-pb -t 7 -r 2000 -z 2000). We parsed the output, in the pairwise mapping format (PAF), to compute the length of the repeat allele in the long-read assembly. Specifically, for each flank, we selected the alignment with the largest number of matching residues ($N_{res}$, column 10 in PAF file), requiring:

1. $N_{res}$>900bp,
2. The length of the target contig containing the matched assembly sequence (column 7 in PAF file) is >10kb, and
3. No competing matches with “Number of residue matches”>0.97*$N_{res}$

We proceeded with analysis if both flanks had an alignment satisfying (1-3). In this case, we further required that

A. The two flanks mapped to the same contig (column 6 in PAF file), and

B. The alignment directions (column 5 in PAF file) of the two flanks were consistent.

If both flanks had alignments satisfying (A-B), we measured the length of the repeat allele in the assembly by computing the distance between the flanking alignments, adjusting for any non-aligned bases at the ends of the flanks (columns 3 and 4 in the PAF file). We finally required that the computed allele length to be greater than –500bp and less than 200*(length of the repeat in GRCh38). Note that we allowed alleles to have negative lengths, possibly reflecting deletions that occurred near the ends of the VNTR or repeat loci whose boundaries were called incorrectly.

We next estimated the number of distinct alleles at each repeat locus (across the subset of the 64 HGSVC2 assemblies for which the algorithm above produced a length measurement). To do so, we counted the number of distinct allele length genotypes, requiring distinct alleles to differ by at least one-quarter of an estimated repeat unit. For repeats with short repeat units, we additionally required distinct alleles to differ by at least 7bp.

To obtain our final list of VNTRs for phasing and imputation optimization, we applied the following filters to the candidate VNTR loci:

1. >50% genotyping rate among HGSVC2 assemblies
2. ≥3 alleles represented among all *N*=64 assemblies
3. ≥2 alleles represented among *N*=12 assemblies from individuals of European descent

Finally, among all loci that satisfied (1-3), we removed regions that had substantial overlap with another VNTR. To do so, we iteratively removed each locus that had substantial overlap with another region (overlap spanning >10% of one of the two regions) where the overlapping region had higher estimated pre-refinement genotyping accuracy (estimated from IBD2 sibling correlation in all SSC participants).

This yielded a filtered set of 15,653 multi-allelic VNTR loci for further analysis.

#### 4 Phasing and imputing VNTR lengths using surrounding SNPs

We performed statistical phasing on WGS read-depth-derived VNTR length estimates ("diploid VNTR content"; see Section 2 above) to estimate haploid allele lengths in SSC participants, which we then imputed from SSC into the UK Biobank cohort based on surrounding SNP-haplotypes. To do so, we adapted the computational algorithm that we previously used to efficiently phase and impute multiallelic protein-coding VNTRs with real-valued length estimates derived from whole-exome sequencing read-depth within UKB (Mukamel et al. 2021). This algorithm is described in detail in Supplementary Text 3 of (Mukamel et al. 2021); in brief, it employs an iterative approach (broadly similar to many algorithms that have been developed for phasing biallelic SNPs) in which haploid allele lengths of each individual in turn are updated according to a probabilistic haplotype-copying model using all other haplotypes as a reference panel, prioritizing copying from haplotypes closely matching the individual's SNP-haplotypes.

To make use of familial relatedness within the SSC cohort and to facilitate imputation from the SSC data set (containing WGS-based SNP calls in hg38 coordinates) to the UK Biobank data set (containing SNP-array genotypes in hg19 coordinates), we made the following minor modifications to our previous phasing and imputation approach. At each VNTR locus:

- We used the IBD maps we generated within SSC families (see above) to identify sib-pairs who inherited the same allele from their mother and inherited the same allele from their father ("IBD2" sibs, which we used for accuracy benchmarks; see below).
- When optimizing parameters for phasing and imputation (using the cross-validation-based procedure described in (Mukamel et al. 2021)), we held out diploid VNTR content estimates for:
    - 400 children (to enable optimization of phasing parameters by maximizing concordance with allele lengths estimated for transmitted parental haplotypes);
    - 400 individuals of European ancestry (to estimate European-ancestry imputation accuracy as described below, holding out full families to prevent relatedness from inflating the benchmark); and
    - 400 individuals of non-European ancestry (again holding out full families).
- For phasing (within SSC), we computed SNP-haplotype similarity based on identity-by-state (IBS) length as described in (Mukamel et al. 2021), which we computed using SNPs with MAF>0.01 in our QC-ed and phased version of the SSC WGS 2 (hg38) variant call set.

- For imputing from SSC into UK Biobank, we computed IBS (at the VNTR's hg19 location) using SNPs with MAF>0.001 that were present in the UKB SNP-array data set (hg19) as well as the SSC data set (lifted from hg38 to hg19).
- For imputation, we restricted VNTR+SNP haplotypes to parents in SSC ($N$=4,688 individuals after sample exclusions; $N$=9,376 haplotypes) to avoid redundancy given the family structure of the SSC cohort. We post-processed the VNTR allele length assigned to each parental haplotype by taking the average of the allele length estimated for that haplotype by our phasing algorithm as well as the allele lengths estimated in any children to which the allele had been transmitted (based on our IBD maps; we restricted to confident transmissions with >2Mb of IBD-sharing).

#### 5 Estimating VNTR genotyping and imputation accuracy and VNTR-SNP linkage disequilibrium

To estimate the accuracy of VNTR length estimates derived from WGS read-depth in individual genomes (before incorporating information from SNP-haplotypes, i.e., “genotype accuracy pre-refinement” in Fig. 1b,c), we used correlations among IBD2 sib-pairs as in our previous work (Mukamel et al. 2021). Explicitly, assuming unbiased error in read-depth-based measurements of diploid VNTR content, we can estimate the accuracy (i.e., $R^2$ vs. truth) of these measurements as:

$$\widehat{R^2}(\text{diploid estimates, truth}) = R(\text{diploid estimate in sib 1, diploid estimate in sib 2}) = \text{"IBD2 }R\text{"}$$

To estimate imputation accuracy, we used a cross-validation-based approach as in our previous work: for 400 individuals held-out from phasing, we imputed VNTR lengths into the held-out individuals and then estimated imputation accuracy as:

$$\widehat{R^2}(\text{imputed estimates, truth}) = \frac{R^2(\text{imputed estimates, held out estimates})}{\text{IBD2 }R}$$

where dividing out by $\text{IBD2 }R$ (an estimate of $R^2(\text{held out estimates, truth})$) accounts for measurement error in the held-out values. To obtain accuracy estimates indicative of imputation performance into the predominantly European-ancestry UK Biobank cohort, we restricted to SSC participants of European ancestry when selecting IBD2 sib-pairs and held-out individuals.

Two potentially counterintuitive features of these accuracy estimates are worth noting:

- Imputation accuracy can sometimes exceed pre-refinement genotype accuracy (i.e., accuracy of the diploid VNTR content measurements on which imputation is based). This behavior typically occurs if a VNTR has a narrow allele length distribution (such that alleles are difficult to distinguish from read-depth) but alleles are well-tagged by nearby SNPs, such that the phasing and imputation model is able to learn which SNP-haplotypes carry which alleles (and use SNPs to predict alleles more accurately than possible from read-depth).
- Imputation accuracy estimates are noisier for VNTRs with lower pre-refinement genotype accuracy (i.e., lower $\text{IBD2 }R$). This behavior is driven by the need to divide by $\text{IBD2 }R$ (which can be a small quantity with sizable uncertainty) when estimating imputation accuracy using cross-validation. While our $\text{IBD2 }R$ estimates typically used ~400 IBD2 sib-pairs at each locus, providing reasonable precision, noise in $\text{IBD2 }R$ occasionally resulted in imputation accuracy estimates that exceeded 1 (presumably due to $\text{IBD2 }R$ having been underestimated by chance).

One caveat of the above benchmarks is that they assume unbiasedness of errors in read-depth-based estimates of diploid VNTR content. We previously observed that exome sequencing coverage depths at VNTRs can be biased by the presence of paralogous sequence variants (PSVs) within repeat units (that can subtly affect exome capture) or by read-mapping biases for very short alleles (Mukamel et al. 2021). While the first issue has much less of an effect on whole-genome sequencing (which does not involve a capture step), to ensure robustness of our results, we performed follow-up analyses of VNTRs of particular interest in which we (i) used a variety of locus-specific techniques to optimize genotyping accuracy (see below); and (ii) validated WGS-derived genotypes against allele lengths directly measured from long-read sequencing data (Supplementary Fig. 1).

##### *Computing VNTR-SNP linkage disequilibrium*

To estimate VNTR-SNP linkage disequilibrium (LD) (Fig. 1c, Supplementary Table 1), we computed the correlation coefficient between "pre-refinement" VNTR genotypes (estimated in individual genomes from WGS depth-of-coverage) and SNP genotypes, and adjusted for the estimated accuracy of VNTR genotypes:

$$\widehat{R^2}(\mathrm{VNTR},\mathrm{SNP}) = \frac{R^2(\mathrm{est.\ prerefinement\ VNTR\ genotypes},\mathrm{SNP\ genotypes})}{\mathrm{IBD2}\ R}.$$

We restricted analysis to 3,904 unrelated SSC participants of European descent. We additionally restricted to SNPs within 500kb of the VNTR, excluded variants within the VNTR, and excluded very rare (MAF<0.0005) variants. For VNTRs at *TMCO1*, *EIF3H*, and *CUL4A*, we additionally estimated VNTR-SNP LD using optimized VNTR genotypes and imp_v3 SNPs dosages in $N$=16,728 UKB participants, obtained by 25x-downsampling the set of 418,136 unrelated, PC-filtered individuals used in our primary analysis. (We did not adjust for VNTR genotype accuracy in UKB). We used these UKB-derived estimates for correlations reported in the main text and to color Manhattan plots (Figs. 3b,c, 4a,b, and 5b,d; Supp. Fig. 4) .

##### *Selection of final VNTR list for imputation into UKB*

We applied the following set of QC filters to select variants suitable for taking forward for imputation into UKB:

1. IBD2R > 0.1 (in SSC)
2. RDIFF > 0.1 (in SSC)
3. FLANKR < 0.5 (in SSC)
4. Imputation $R^2$ > 0.1 (in SSC participants of European descent)
5. We excluded variants with the major histocompatibility comples (MHC) locus (chr6:29mb-33mb)

This resulted in the final set of 9,561 multiallelic VNTR loci for analysis in UKB.

#### 6 Optimizing genotyping of VNTRs of particular interest

For each VNTR for which our association analysis and fine-mapping pipeline identified a potentially-causal phenotype association of particular interest (specifically, associations with disease traits and associations of *CUL4A* with erythrocyte traits), we performed follow-up analyses to optimize accuracy of VNTR allele length estimates and verify robustness of results. We did so by analyzing WGS data subsequently released for *N*=200K UKB participants (Halldorsson et al. 2022), which we then used as a reference panel for imputation into the remainder of the UK Biobank cohort. Beyond the increased phasing and imputation accuracy afforded by the much larger size of this reference panel (compared to our initial analysis of *N*=8,936 SSC participants), we also obtained further improvements in VNTR genotyping accuracy by developing statistical models tailored to the allele distribution at each locus. Specifically, we incorporated information from 151bp reads that spanned short VNTR alleles (at *TMCO1* and *EIF3H*), and we optimized the selection of reads counted in read-depth-based measurements of VNTR length.

##### *TMCO1*

<u>*Improved TMCO1 VNTR genotyping by combining spanning-read and read-depth information.*</u>

The *TMCO1* VNTR has a bimodal allele length distribution, with the 1-repeat allele having high frequency (>0.85) in all continental populations, alleles containing 2 to 4 repeats being very rare, and expanded alleles with ≥5 repeats comprising the remainder of the allele distribution (Fig. 3a,d). The repeat unit length of 28bp meant that *TMCO1* VNTR alleles with 1 to 4 repeats were consistently spanned by multiple 151bp reads indicating their presence. (We also searched for evidence of 0-alleles but did not find any evidence that such alleles existed.) While expanded alleles with ≥5 repeats could not be distinguished by single reads, the presence of such an allele could easily be detected based on observations of 151bp reads that partially overlapped the VNTR, and additionally, the lengths of expanded alleles could be estimated by counting the number of reads internal to the VNTR (similar to the read-depth-based strategy we used in initial genotyping, but greatly reducing noise by restricting to within-VNTR reads).

We therefore implemented a hybrid genotyping strategy (similar to the approach we previously used to genotype *TENT5A* alleles from WES (Mukamel et al. 2021)) that combined direct read-level information (used to identify short alleles and to detect the presence of expanded alleles) with read-depth information (used to estimate the lengths of longer alleles). Specifically, for each individual, we applied the following procedure:

- Identify the minimum- and maximum-length allele indicated by direct read-level evidence (which could be the same allele, indicating a homozygote).
- If the maximum-length allele indicated has length ≤4, set the individual's (unphased) genotype to be the minimum-length and maximum-length allele.

- Otherwise:
  - If the minimum-length allele has length ≤4 (i.e., the individual is heterozygous for an expanded allele), then estimate the number of repeats in the expanded allele as:
    5 + (# within-VNTR reads) / (# reads in $\pm$5kb flanks) x (calibration factor).
  - Otherwise, estimate the total number of repeats in the two expanded alleles as:
    10 + (# within-VNTR reads) / (# reads in $\pm$5kb flanks) x (calibration factor).

Based on empirical analyses of WGS data from SSC, UKB, 1000 Genomes 30x, and GTEx, the calibration factor above that is required to convert read counts to absolute estimates of expanded allele lengths appeared to be data set-specific. We therefore estimated this calibration factor independently for each data set in which we performed analysis using the following approach:

- First, we estimated the calibration factor in the 1000 Genomes 30x data set (Byrska-Bishop et al. 2022) by identifying 17 heterozygous carriers of expanded alleles with lengths that could be exactly determined from a long-read assembly of either the carrier or a related individual included in HGSVC2 (Ebert et al. 2021) or HPRC (Liao et al. 2022). We set the calibration factor for 1000 Genomes 30x to the value that caused the mean estimated length of expanded alleles in these 17 individuals to equal the mean of the exact long-read-derived lengths.
- Next, we estimated mean lengths of expanded alleles in heterozygous carriers in each 1000 Genomes Project continental population (Fig. 3a) by applying the calibration factor estimated above to all samples in the 1000 Genomes 30x WGS data set.
- Finally, for each other WGS data set we analyzed (all of which were predominantly EUR-ancestry), we set the calibration factor to the value that caused the mean estimated length of expanded alleles in heterozygous carriers to match the mean expanded allele length we estimated in the previous step for 1000 Genomes EUR participants.

*Optimized phasing and imputation of TMCO1 VNTR genotypes.*

For each whole-genome-sequenced individual, the above strategy produced a pair of (unphased) allele length estimates with the property that calls of short alleles (≤4 repeats; usually the 1-allele) were discrete and nearly always correct, and detection of expanded alleles (≥5 repeats) was also nearly always correct, but lengths of expanded alleles were only approximately measured (by read-counting). We next needed to phase these estimates onto SNP-haplotypes in order to denoise estimated lengths of expanded alleles (by averaging estimates across individuals with long shared SNP-haplotypes) and to enable imputation into SNP-haplotypes of unsequenced UKB participants. While we could do so using our standard phasing and imputation algorithm (which treated all genotype estimates as continuous, real-valued measurements), the discrete information available here from read-level analysis allowed a simpler, more accurate approach.

To phase each individual's pair of allele length estimates onto the individual's SNP-haplotypes and refine estimates of expanded allele lengths, we did the following:

1. Determine which of the individual's two SNP-haplotypes carries the shorter allele and which SNP-haplotype carries the longer allele. We did so by counting, for each of the target individual's two SNP-haplotypes, how many of the carriers of the top 20 longest SNP-haplotype-matches had read-level support for the target individual's longer allele. We then assigned the target individual's longer allele to the SNP-haplotype with more "votes" from top haplotype matches.
2. Refine the length estimate of each detected expanded allele by taking a weighted average of the allele length estimated in the target individual together with allele lengths estimated in individuals who (i) shared a long SNP-haplotype with the target allele; and (ii) carried exactly one expanded allele (presumably on the shared haplotype). We computed this weighted average using the haplotype-copying probabilities we used in our previous work (Mukamel et al. 2021), which are a function of IBS-sharing length and three tunable parameters ($K_{\text{top}}$, $\ell_0$, and $p_{\text{reg}}$). We tuned these parameters using a grid search that utilized cross-validation in IBD2 sib-pairs heterozygous for an expanded allele. Specifically, we held out one member of each sib-pair and chose the parameter combination that maximized correlation between held-out estimates of expanded allele lengths and refined estimates of expanded allele lengths in the non-held-out siblings.

To impute VNTR allele lengths into unsequenced individuals, we used the same haplotype-copying model but re-optimized the three parameters to maximize imputation accuracy in cross-validation (using 400 held-out samples).

*Validating accuracy of TMCO1 VNTR allele length estimates.*

To verify the accuracy of our genotyping strategy at *TMCO1*, we compared VNTR allele lengths we estimated in 1000 Genomes 30x WGS (after phasing together with allele lengths we estimated in UKB *N*=200K WGS) to allele lengths derived from long-read assemblies of HGSVC2 samples (summing across each individual's two alleles). This comparison demonstrated high accuracy ($R^2 = 0.99$; Supplementary Fig. 1a).

*Estimating TMCO1 VNTR allele lengths in GTEx.*

To estimate unphased TMCO1 allele lengths in GTEx, we used the same strategy of combining spanning-read and read-depth information that we used to analyze the UKB *N*=200K WGS and 1000 Genomes 30x WGS data, with just one minor difference that arose from 58 GTEx samples having been sequenced using 100bp reads instead of

151bp reads. We could still use read-level information to determine which of these individuals carried expanded (≥5-repeat) alleles, but we did not attempt to use within-VNTR read counts to estimate the lengths of these expanded alleles, instead setting their initial length estimates to the mean expanded allele length. We then phased the allele length estimates in GTEx together with allele lengths estimated in UKB N=200K WGS and 1000 Genomes 30x WGS to maximize accuracy.

##### *EIF3H*

<u>*Improved EIF3H VNTR genotyping by modeling read-level information.*</u>

Most *EIF3H* VNTR alleles contain 2 to 6 repeats of a 27bp unit followed by a partial repeat unit (13bp). Consequently, alleles with ≤4 full repeats could usually be detected from spanning 151bp reads in the UKB *N*=200K WGS data set. Alleles with ≥5 repeats were too long to genotype from spanning reads, but reads that partially overlapped the VNTR could be informative of the presence of a ≥5-repeat allele, and reads internal to the VNTR indicated the presence of a ≥6-repeat allele. Altogether, read-level information was thus usually sufficient to deduce a confident (unphased) genotype call in a given individual. However, synthesizing all of this information while accounting for occasional false-positives (i.e., observations of reads putatively supporting an allele that is not actually present) and false-negatives (i.e., absence of observations of reads supporting an allele that is present) was not straightforward, as we needed to consider how to weigh evidence from counts of reads in seven different categories:

- span1, span2, span3, span4 (i.e., reads spanning VNTR alleles with 1-4 full repeat units)
- flank4+, flank5+ (i.e., reads partially overlapping the VNTR indicating ≥4 or ≥5 repeats)
- internal (i.e., reads completely within the VNTR indicating ≥6 repeats).

We therefore developed a Bayesian genotyping strategy based on a generative model in which we assumed reads from each of the seven categories were generated independently (conditional on an individual's genotype). Letting $\mathrm{CN1}, \mathrm{CN2}$ denote the numbers of full repeat copies on the individual's two haplotypes, we assumed:

$$P(\,\mathrm{CN1}, \mathrm{CN2} \mid \mathrm{obs.\,reads}\,) \propto P(\mathrm{CN1})\,P(\mathrm{CN2}) \prod\nolimits_{\mathrm{category}} P(\#\ \mathrm{reads\ in\ category} \mid \mathrm{CN1}, \mathrm{CN2})$$

where for each of the seven categories of reads, we modeled $P(\#\ \mathrm{reads\ in\ category} \mid \mathrm{CN1}, \mathrm{CN2})$ using a Poisson distribution with

$$\lambda = \lambda_{\#(\mathrm{CN1},\mathrm{CN2\ contributing\ to\ category})} \cdot (\mathrm{local\ read\ depth\ in\ 5kb\ flanks})$$

where the rate parameters $\lambda_0, \lambda_1, \lambda_2$ are defined as follows:

- $\lambda_0$ (neither $\mathrm{CN1}$ nor $\mathrm{CN2}$ should generate reads in the category): estimate based on empirical frequency of observing false-positive reads (in samples with strong evidence that they carry only alleles that should not produce reads in the category)

- $\lambda_1$ (exactly one of the two alleles generates reads in the category): estimate based on empirical frequency of observed reads in samples with good evidence that they carry exactly one such allele
- $\lambda_2 = 2\lambda_1$ (both alleles generate reads in the category, so twice as many reads are expected).

After estimating $\lambda_0, \lambda_1, \lambda_2$ as indicated above, we then used an expectation-maximization (EM) algorithm to estimate the frequencies of alleles with 1 to 6 repeat units to use as priors $P(\text{CN1}), P(\text{CN2})$. (We did not observe evidence of 0-repeat alleles, and while analysis of within-VNTR read counts indicated that rare 7-repeat alleles also exist, they are sufficiently rare that modeling them distinctly from 6-alleles was not necessary.)

*Optimized phasing and imputation of EIF3H VNTR genotype probabilities.*

For each whole-genome-sequenced individual, the above algorithm produced posterior probabilities for each possible genotype $\{\text{CN1}, \text{CN2}\}$ with no information about phase. For most individuals, a single genotype was by far the most likely (with only the phase of the alleles being unknown), but for some individuals, multiple genotypes had similar posterior probabilities. We therefore leveraged information from shared SNP-haplotypes to help resolve uncertain genotypes and to phase each individual's pair of alleles onto the individual's SNP-haplotypes. We did so by running four iterations of the following algorithm, applied to each individual in turn:

- For each of the individual's two SNP-haplotypes, count how many of the five longest SNP-haplotype matches are believed to carry a 1-allele, 2-allele, …, 6-allele (adding a pseudocount of 0.5 for each allele).
- Adjust the likelihood of each $\{\text{CN1}, \text{CN2}\}$ genotype by multiplying by the relevant numbers of votes of support from SNP-haplotype-matches.
- Select the $\{\text{CN1}, \text{CN2}\}$ genotype with highest adjusted likelihood.
- Set the phase of the shorter/longer allele to match the shorter/longer of the mean allele length estimated in the five best matches for each SNP-haplotype.

We imputed VNTR allele lengths into unsequenced individuals using the same approach as at *TMCO1* (again optimizing imputation parameters via cross-validation in 400 held-out samples).

*Validating accuracy of EIF3H VNTR genotypes.*

To verify the accuracy of our genotyping strategy at *EIF3H*, we compared VNTR genotypes we estimated in 1000 Genomes 30x WGS (after phasing within this cohort) to allele lengths derived from long read assemblies of HGSVC2 samples (summing across each individual's two alleles). This comparison demonstrated high accuracy ($R^2 = 0.99$; Supplementary Fig. 1b).

##### *CUL4A*

*Estimating CUL4A VNTR allele lengths from WGS read-depth.*

To efficiently estimate diploid VNTR content at *CUL4A* in the *N*=200K UKB WGS data release, we counted reads aligning fully within the VNTR region in GRCh38 as well as in 10kb flanks on each side (restricting to reads with SAM flags 0x53, 0x63, 0x93, 0xA3, 0x51, 0x61, 0x91, or 0xA1). The count of flanking reads served as an approximate measure of local sequencing coverage for each sample, allowing us to estimate VNTR allele length (up to a constant calibration factor; see below) as the ratio of the number of within-VNTR reads to the number of flanking reads. To account for the possibility of copy-number variants influencing flanking read counts in a small fraction of samples, we excluded samples with outlier flank read counts (>2.5 s.d. from the mean on a log scale). We then phased these length estimates and imputed into the remainder of the UKB cohort using the same approach as in our previous analysis of UKB *N*=50K WES data (Mukamel et al. 2021).

*Calibrating CUL4A VNTR allele length estimates.*

The above pipeline produced unscaled allele length estimates that were not calibrated to absolute (base pair) lengths. We therefore calibrated *CUL4A* allele length estimates derived from WGS read-depth in UKB by imputing allele lengths from UKB into SNP-haplotypes for 1000 Genomes Project participants (Byrska-Bishop et al. 2022) and calibrating against allele lengths derived from long-read assemblies in the HGSVC2 data set (Ebert et al. 2021). Specifically, we estimated a single scaling factor by regressing long-read-derived allele lengths on WGS-read-depth-derived (imputed) estimates (summed across each individual's two alleles), setting the intercept to 300bp (because only VNTR alleles >150bp can produce 151bp reads that align fully within the VNTR region in GRCh38). We performed this regression using the six EUR individuals included in HGSVC2 (because imputation accuracy was highest in EUR).

*Validating accuracy of CUL4A VNTR allele lengths derived from WGS read-depth.*

Separately, to verify the accuracy of our WGS read-depth-based approach to measuring *CUL4A* VNTR allele lengths, we subsequently ran the same read-counting pipeline directly on WGS read alignments in the 1000 Genomes 30x data set (Byrska-Bishop et al. 2022). We then compared these diploid VNTR content estimates to allele lengths derived from long read assemblies of HGSVC2 samples (summing across each individual's two alleles), observing high concordance ($R^2 = 0.97$; Supplementary Fig. 1c).

##### *CHMP1A, INS, and METRNL*

Similar to *CUL4A*, we estimated diploid VNTR content at *CHMP1A*, *INS*, and *METRNL* in *N*=200K UKB WGS data by counting reads aligning fully within the VNTR region and dividing by the count of reads aligning to the 10kb flanks on each side. (For each of these four VNTRs, nearly all alleles are >150bp, so counting reads aligning fully within the VNTR – i.e., excluding reads that span its left or right edges – reduces noise.) We again excluded samples with particularly low or high counts of reads aligning to the 10kb flanks, restricting to the middle 95% of the distribution (i.e., excluding samples in the top or bottom 2.5%). We then phased and imputed into the remainder of the UKB cohort as before.

Rerunning the association and fine-mapping analysis using the updated allele length estimates increased confidence in causality for the associations of the *CHMP1A* and *INS* VNTRs with hypertension and type 1 diabetes (FINEMAP posterior probability = 1.00 and 0.91, respectively) but decreased confidence in causality of the association of the *METRNL* VNTR with cataracts (FINEMAP posterior probability = 0.03). These results are reported in Supplementary Table 4.

#### 7 Phenotype refinement for disease-associated VNTRs

For the two strongest disease associations we observed, involving VNTRs at *TMCO1* and *EIF3H*, we sought to bolster the statistical evidence of association by: 1) refining the associated disease phenotypes via ICD-10 subcategories; and 2) curating additional, related phenotypes not included in the original set of 786 phenotypes we tested for association.

##### *Glaucoma*

We sought to increase power and statistical resolution to interrogate the relationship between the VNTR at *TMCO1* and glaucoma by refining the associated glaucoma phenotype. We initially observed a strong association between the VNTR at *TMCO1* and the glaucoma phenotype curated by UKB, categorized under the ICD-10 code H40. SNPs at *TMCO1* in LD with the VNTR had previously been associated with primary open-angle glaucoma (POAG) (Burdon et al. 2011). A substantial fraction of glaucoma cases in UKB are classified as primary angle-closure glaucoma (PACG), a disease that has little etiological overlap with POAG (Wiggs and Pasquale 2017). Therefore, we sought to remove known PACG (ICD-10 code H40.2) from the disease phenotype. To do so, we extracted the ICD-10 codes recorded for diagnoses made during hospital inpatient admissions (UKB data field 41270, accessed via the Research Analysis Platform (RAP) on 06/03/2022). We then curated a new binary glaucoma phenotype, where we included as cases all participants with a glaucoma diagnosis (either in the original UKB-curated phenotype, or a H40 code present in data field 41270), and then removed all participants with a specific diagnosis of PACG (H40.2). Individuals with diagnoses of both POAG (H40.1) and PACG (H40.2) were considered as cases. Among the PC-filtered, unrelated set of 418,136 UKB participants in our primary analysis, we identified a total of 15,334 glaucoma cases, 1,216 of which were classified as PACG (and not POAG), leaving 14,118 cases in our final analysis. We used the resulting glaucoma phenotype for all follow-up analyses, with the exception of the estimation of the overall disease burden of expanded *TMCO1* VNTR alleles, for which we used explicit diagnoses of POAG (H40.1).

##### *Intraocular pressure*

We sought independent statistical evidence of the *TMCO1* VNTR's association with glaucoma by analysis of intraocular pressure (IOP), a major risk factor for glaucoma that was measured in ~130K UKB participants but was not in our initial analysis set. We curated a phenotype derived from IOP measurements following the practices of a recent IOP GWAS performed using UKB data (Khawaja et al. 2018). We extracted UKB data fields 5254 and 5262, which recorded measurements of corneal-compensated IOP in the left and right eyes, respectively. Each participant had up to two measurements taken from each eye. We removed outlier measurements (<7 and >30 mmHg, approximately ~1% of all measurements), and averaged the remaining measurements for each participant. We used the resulting IOP phenotype for all association analyses.

To assess the effects of specific *TMCO1* VNTR alleles (Fig. 3e), we normalized the resulting IOP phenotype by regressing out age, $age^2$, and sex, and applying a linear transformation to obtain a distribution with mean 0 and standard deviation 1. To minimize confounding from IOP-lowering drugs administered to glaucoma patients, and to ensure the IOP association we observed was statistically independent of the glaucoma association, we excluded all participants with a glaucoma diagnosis from all IOP analyses.

##### *Colon polyps*

We sought to increase power and statistical resolution to interrogate the relationship between the VNTR at *EIF3H* by refining the associated phenotype categorized under ICD-10 code K63 (other diseases of the intestine). We extracted the ICD-10 codes recorded from hospital inpatient admissions (UKB data field 41270, accessed via the RAP on 06/03/2022). The majority (77%) of K63 reports were subclassified as K63.5 (colon polyps), and association analyses revealed that K63.5 was the only K63 subcategory that was significantly associated with the *EIF3H* VNTR length. In our final analyses, we analyzed a binary phenotype where cases included only individuals with specific ICD-10 reports of K63.5 in data field 41270 (22,715 cases among the PC-filtered, unrelated set of 418,136 UKB participants in our primary analysis).

##### *Colorectal cancer*

Given the strong association between the *EIF3H* VNTR and colon polyps, and previous reports that SNPs near *EIF3H* strongly associated with colorectal cancer (CRC), we hypothesized that the *EIF3H* VNTR might also associate with CRC. We sought to test this hypothesis in UKB by direct analysis of CRC, a phenotype not included in our original list of 786 phenotypes tested for association. We extracted the ICD-10 codes obtained from UK cancer registries (UKB data field 40006 with 17 instances, accessed via RAP on 06/03/2022). We identified 6,824 participants (out of 418,136 PC-filtered unrelated individuals) with reports of colorectal cancer (ICD-10 codes C18, C19 or C20). Of these CRC cases, $N$=1,988 participants also had a K63.5 diagnosis.
