## Supplementary Figures for "Repeat polymorphisms in non-coding DNA underlie top genetic risk loci for glaucoma and colorectal cancer"

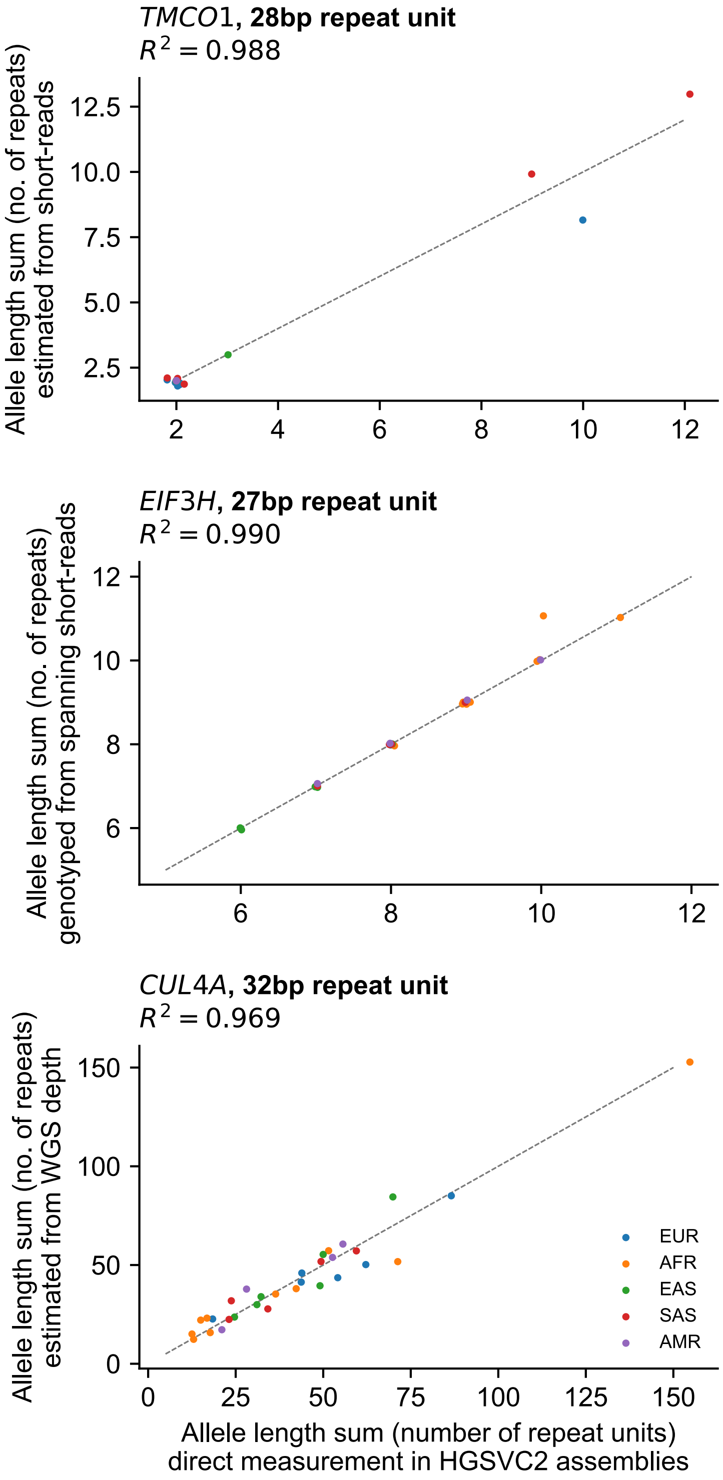


**Supplementary Figure 1. VNTR allele lengths estimated from short-read WGS at *TMCO1*, *EIF3H*, and *CUL4A* are highly concordant with direct measurements in HGSVC2 long-read assemblies.** Scatter of allele lengths estimated from WGS (summed across each individual's two alleles) vs. lengths measured directly in *N*=64 HGSVC2 haploid long-read assemblies (*N*=32 diploid individuals) for VNTRs at *TMCO1* (top), *EIF3H* (middle), and *CUL4A* (bottom). The strategies for genotyping VNTR alleles from WGS were locus-specific and utilized particular features of each allele distribution as well as SNP-haplotype modeling (for *TMCO1* and *EIF3H*; Supplementary Note). Markers are plotted with a small amount of jitter so that all data is visible. Marker color, continental ancestry.

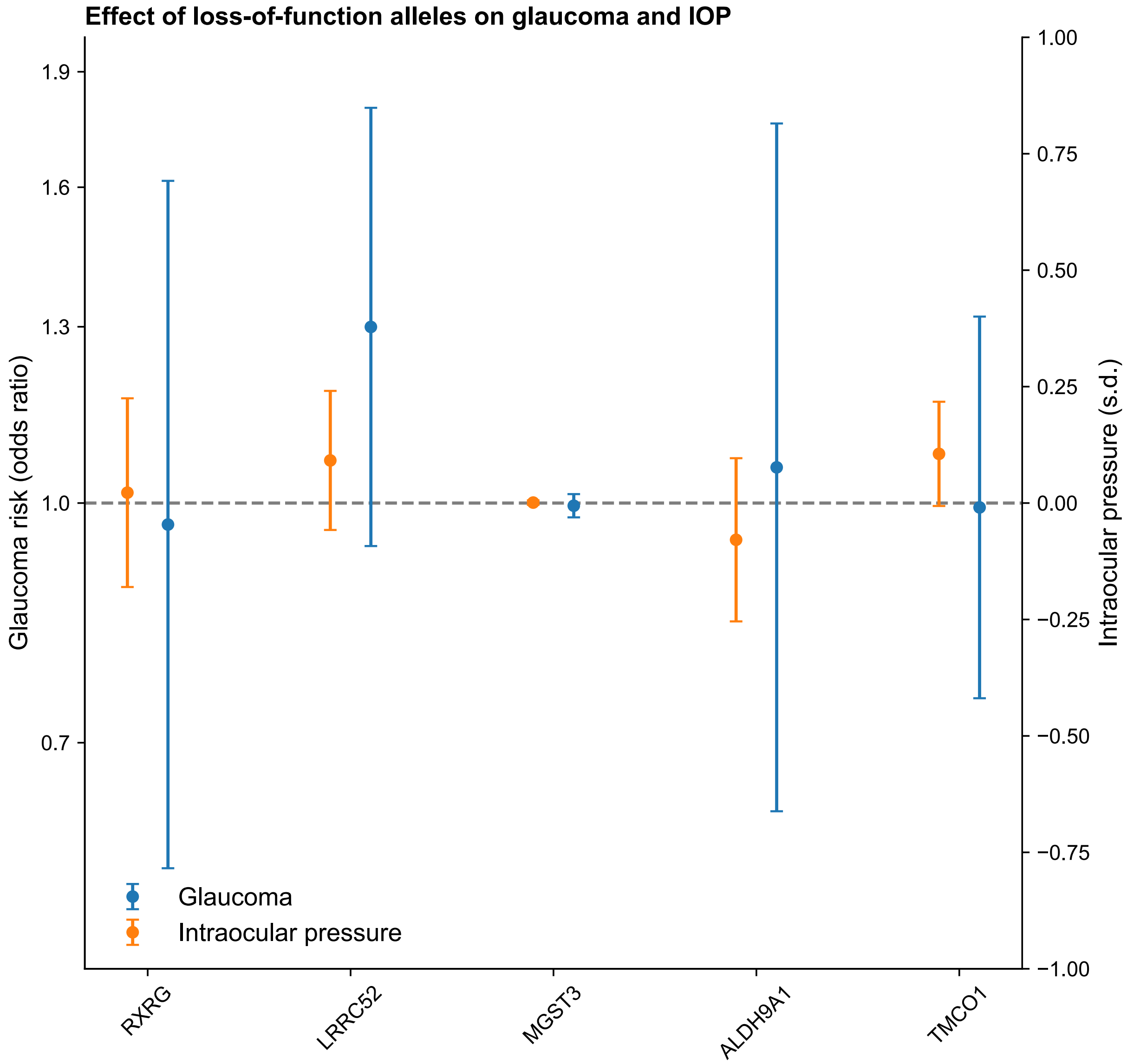


**Supplementary Figure 2. Protein-truncating variants in *TMCO1* and nearby genes do not appear to influence glaucoma risk or intraocular pressure.** Loss-of-function (LOF) effect sizes for glaucoma risk (blue) and intraocular pressure (orange) for each gene within 1Mb of *TMCO1* for which >100 UKB participants of European ancestry carried a protein-truncating SNP or indel variant predicted to cause LOF (Backman et al. 2021). Error bars, standard errors.

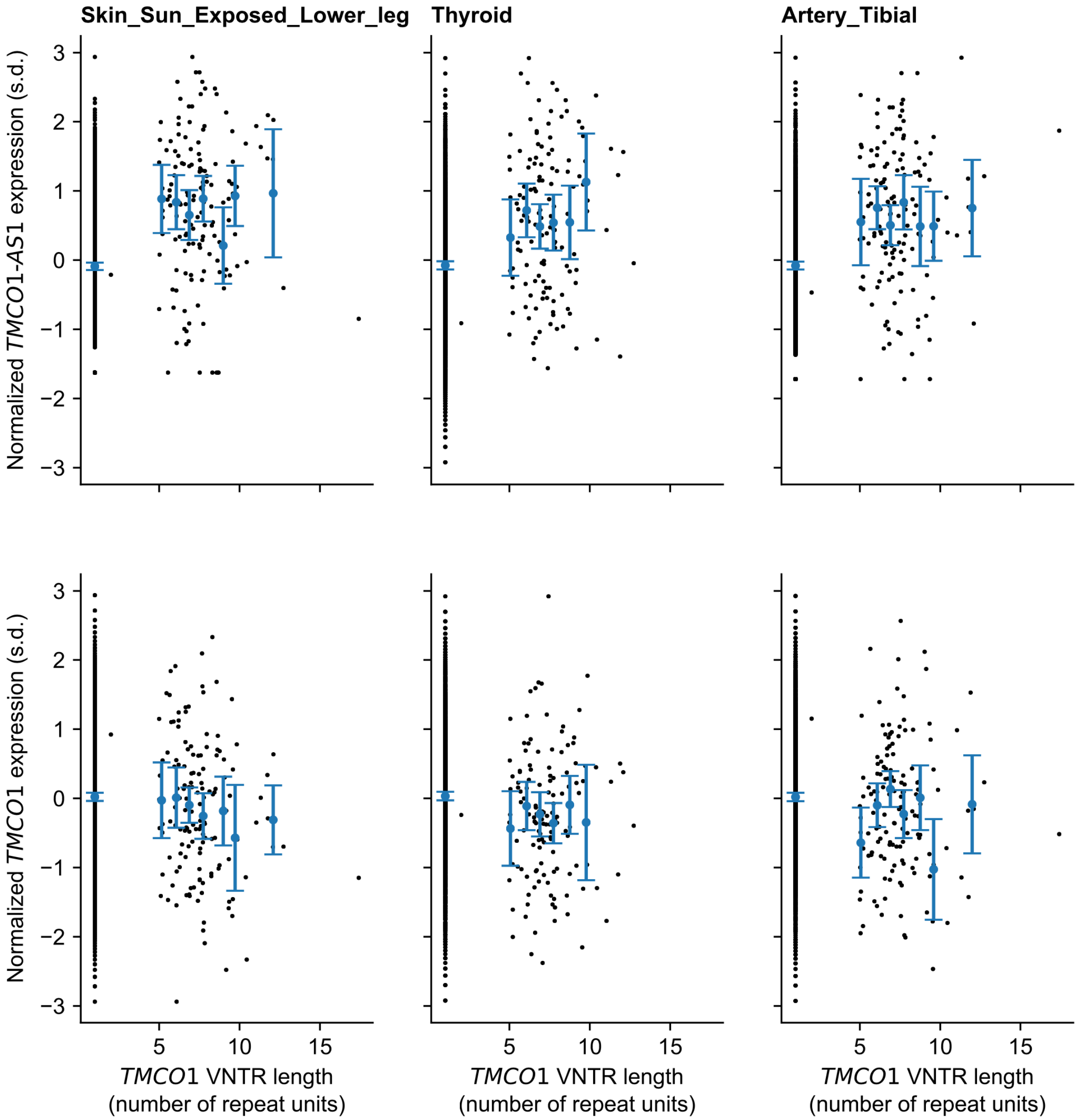


**Supplementary Figure 3. Associations between *TMCO1* VNTR length and nearby expression show no evidence of an allelic series.** Scatter (black dots) of normalized *TMCO1-AS1* expression (top) and *TMCO1* expression (bottom) vs. *TMCO1* VNTR length in the three tissues exhibiting the strongest VNTR association with expression: sun-exposed skin (left), thyroid (middle), and tibial artery (right). Because normalized expression measurements were not allele-specific, each sample is plotted twice (with same y-axis value), once for each allele (x-axis). Larger markers, mean expression across carriers of each VNTR allele; error bars, 95% CIs.

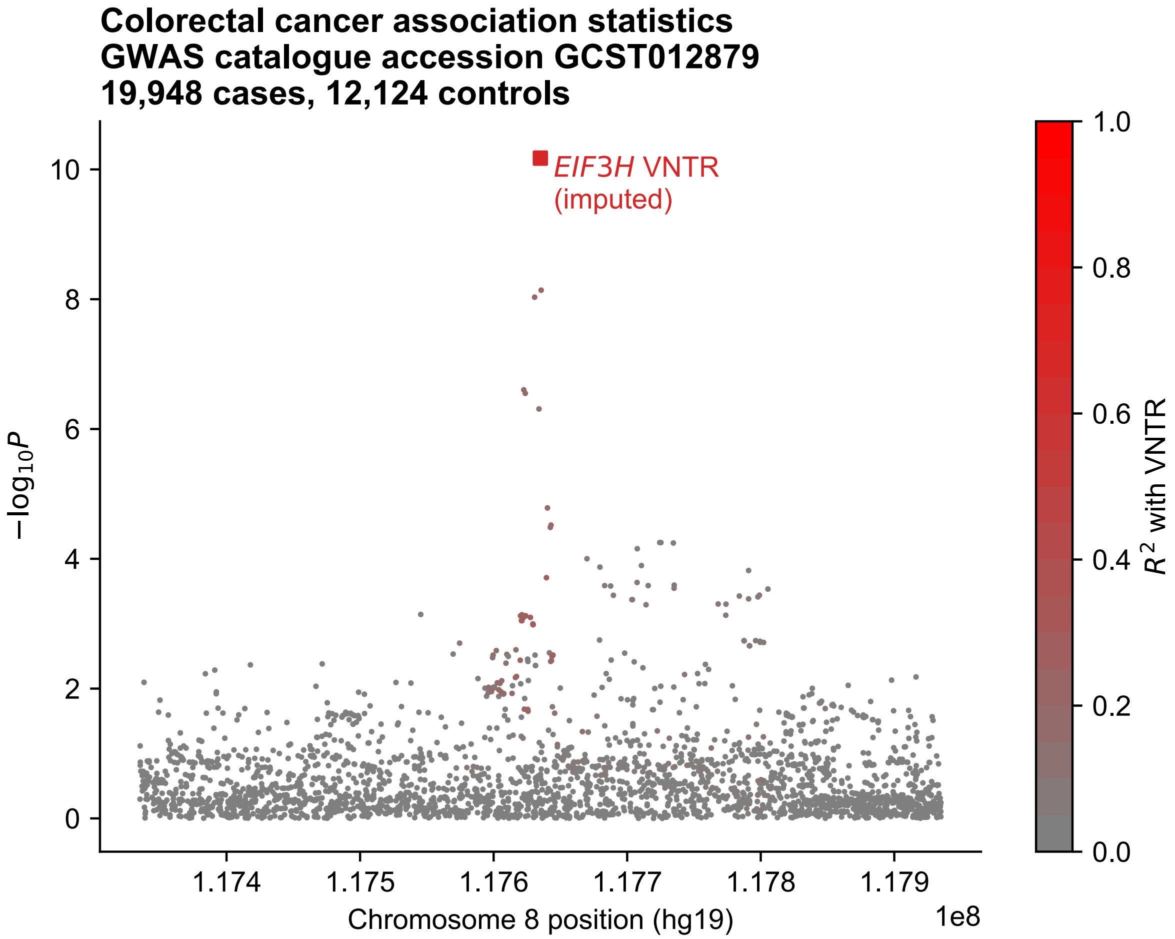


**Supplementary Figure 4. Imputed VNTR association with colorectal cancer is stronger than reported associations to SNPs at *EIF3H,* in GWAS data independent of that in Fig. 4.** Manhattan plot of imputed VNTR association (Methods) and reported SNP associations with colorectal cancer risk at *EIF3H*. Marker color, linkage disequilibrium with VNTR estimated in UKB.

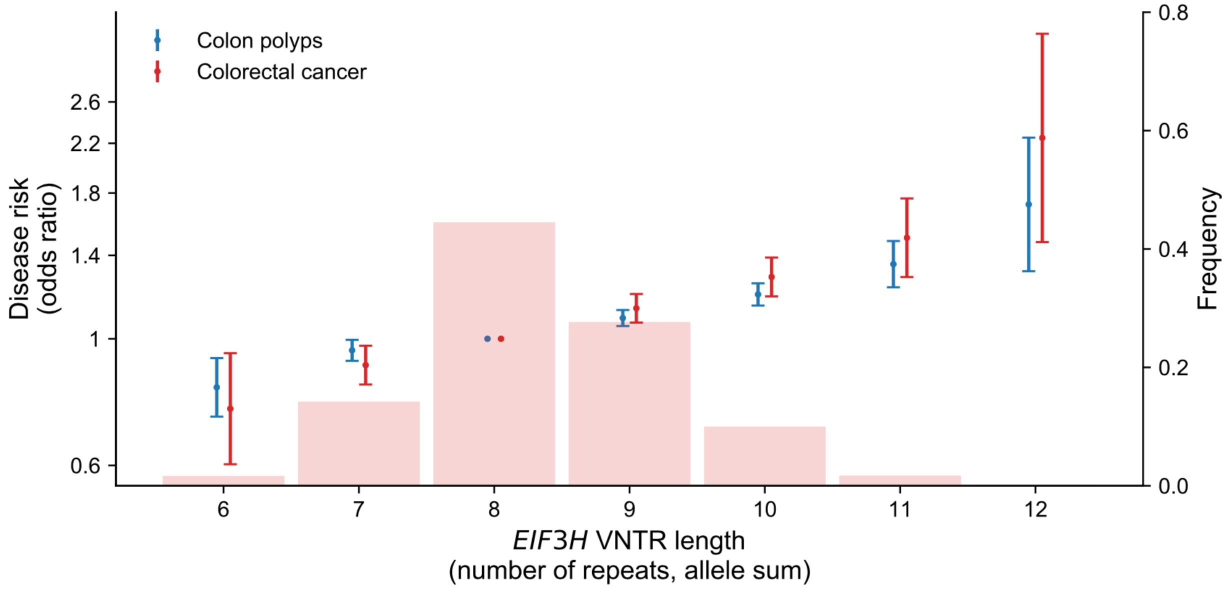


**Supplementary Figure 5. Common repeat length polymorphism downstream of *EIF3H* appears to produce a >2-fold range of colorectal cancer risk in individuals of European ancestry.** Frequencies of "diploid" VNTR allele lengths (summed across each individual's two alleles) observed in European-ancestry UKB participants (histogram, right axis) and their effect sizes (markers, left axis) for colorectal cancer (red) and colon polyps (blue). Odds ratios are computed from logistic regression coefficients, with sex, age, age$^2$, and 20 PCs included as covariates. Error bars, 95% CIs.

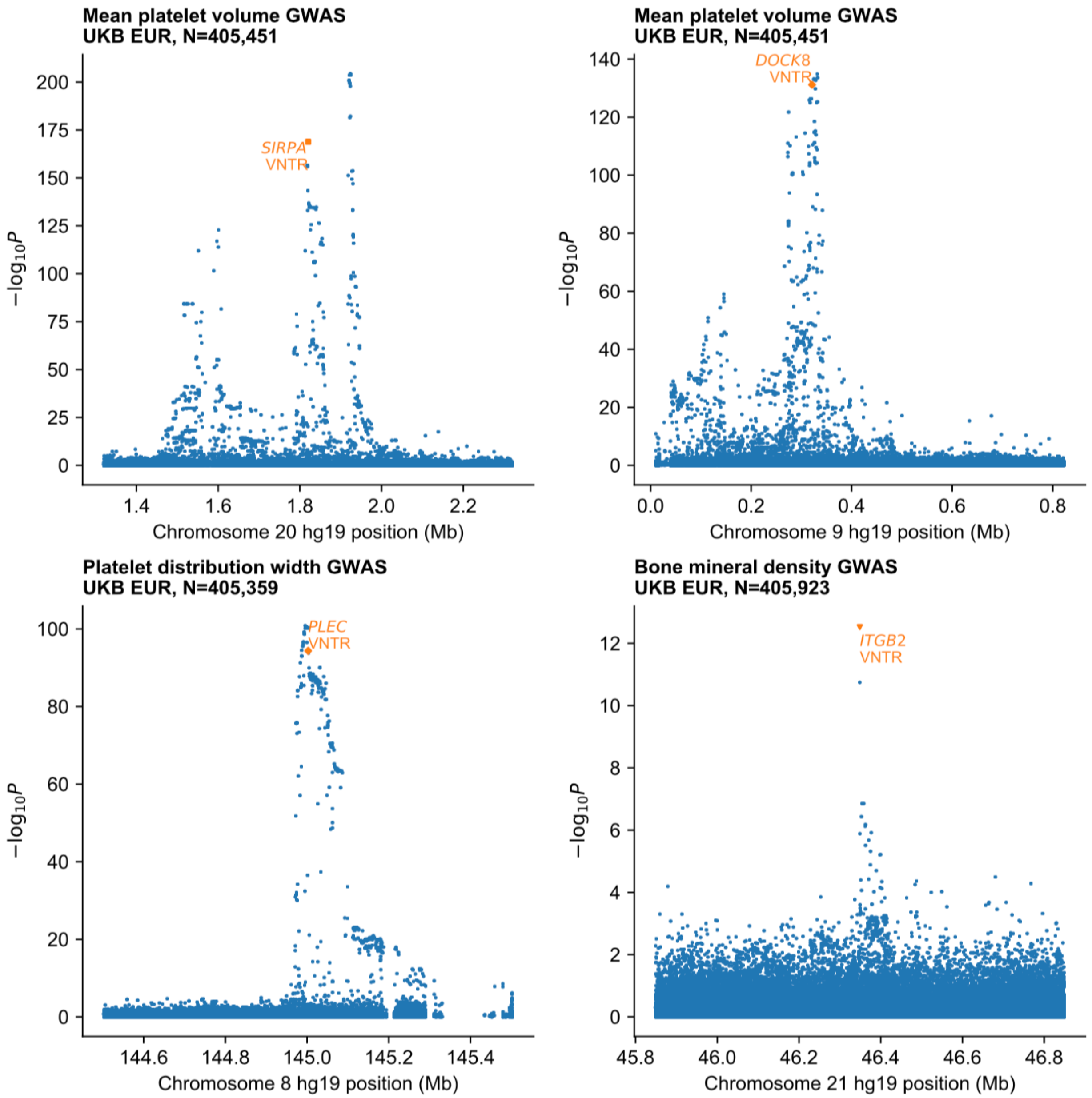


**Supplementary Figure 6. Imputed VNTR lengths at *SIRPA, DOCK8, PLEC* and *ITGB2* associated with phenotypes in UKB.** Manhattan plots displaying VNTR and SNP association statistics for associations at *SIRPA* (top left), *DOCK8* (top right), and *PLEC* (bottom left) with platelet traits, and at *ITGB2* (bottom right) with bone mineral density. At each of these loci, FINEMAP assigned a high posterior probability of causality to the VNTR (PIP>0.99).

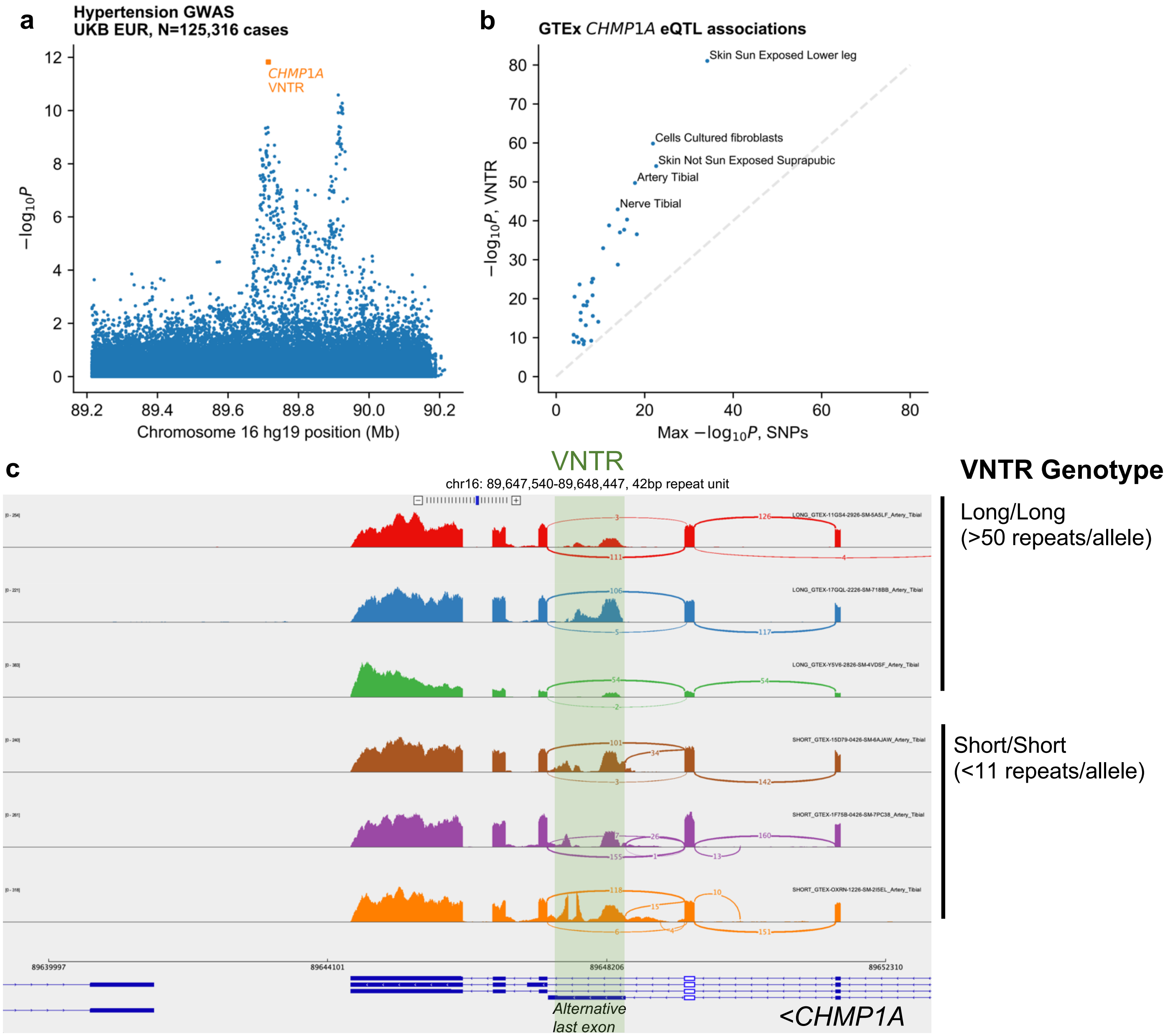


**Supplementary Figure 7. Length of a VNTR within *CHMP1A* associates with hypertension and *CHMP1A* expression. a)** Manhattan plot of hypertension (UKB field 131286, categorized under ICD-10 code I10) associations at *CHMP1A*. **b)** Scatter of VNTR vs. strongest SNP association strength with normalized *CHMP1A* expression in each of 34 tissues for which the VNTR association reached study-wide significance ($P$<5 x $10^{-9}$). In each tissue, longer VNTR alleles associated with decreased *CHMP1A* expression. **b)** Sashimi plots (IGV screenshot) of aligned reads for 6 GTEx tibial artery biosamples, 3 from individuals with homozygous long VNTR genotypes (top), and 3 from individuals with homozygous short VNTR genotypes (bottom). Short VNTR alleles appeared to associate with increased usage of an alternative, VNTR-spanning last exon. Counts of reads supporting splice junctions for introns emanating from the exon colored white (chr16:89,649,350-89,649,496) are displayed. Green highlight, location of VNTR.

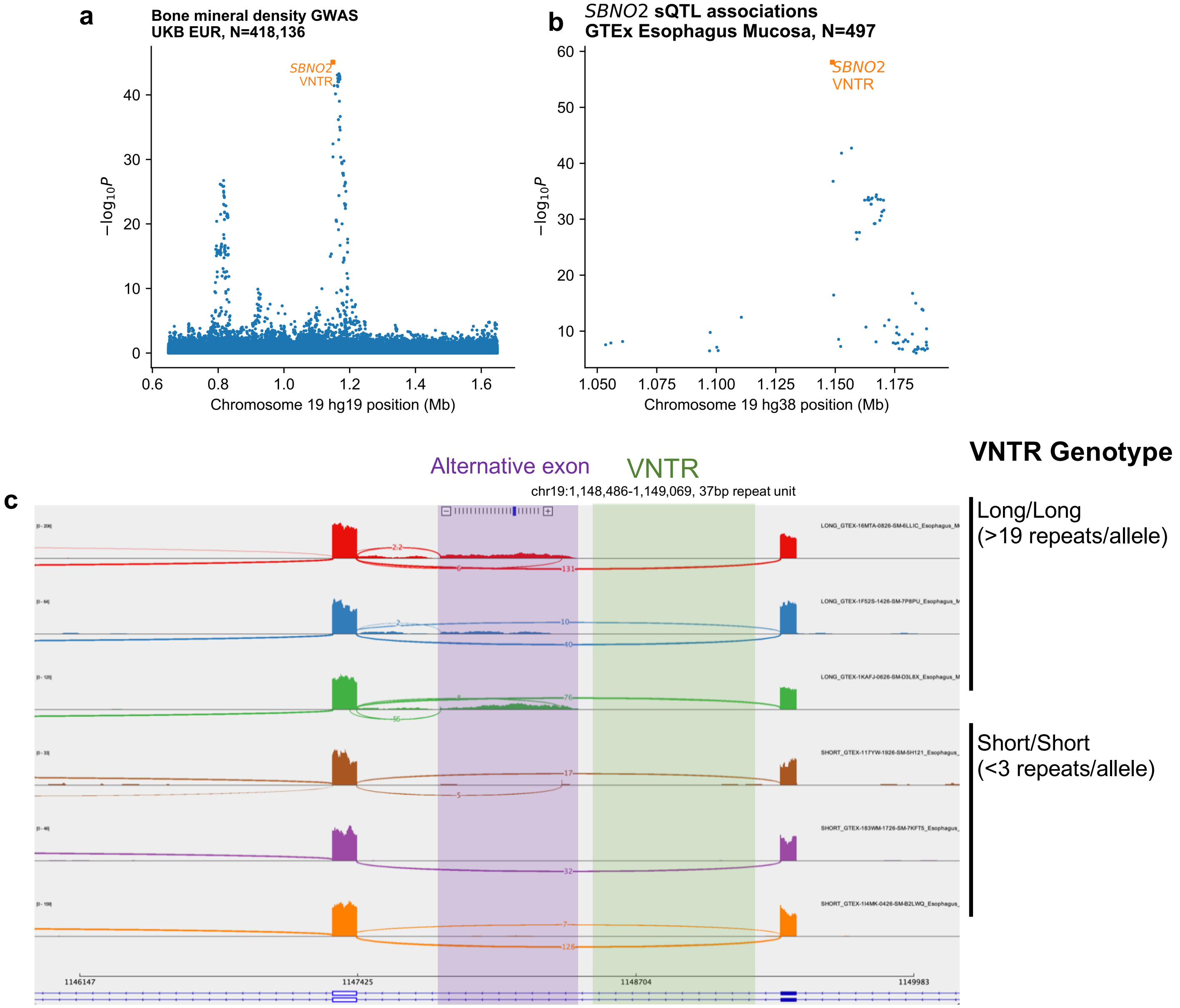


**Supplementary Figure 8. Length of a VNTR within *SBNO2* associates with bone mineral density and gene splicing. a,b)** Manhattan plots of SNP and VNTR associations with bone mineral density (a) and *SBNO2* splicing (b). The splicing phenotype is derived from the excision ratio for the VNTR-spanning intron chr19:1,147,420-1,149,369 in esophagus mucosa samples. SNP associations are displayed for all significantly associated SNPs reported by GTEx. **c)** Sashimi plots (IGV screenshot) of aligned reads for 6 GTEx esophagus mucosa biosamples, 3 from individuals with homozygous long VNTR genotypes (top), and 3 from individuals with homozygous short VNTR genotypes (bottom). Long VNTR alleles appeared to associate with increased usage of an unannotated exon (purple highlight) immediately downstream of the VNTR (green highlight). Counts of reads supporting splice junctions for introns emanating from the exon colored white (chr19:1,147,309-1,147,420) are displayed.
